# Major depression, suicidal behaviors and neuroticism are pro-atherogenic states driven by lowered reverse cholesterol transport

**DOI:** 10.1101/2023.02.10.23285746

**Authors:** Ketsupar Jirakran, Asara Vasupanrajit, Chavit Tunvirachaisakul, Marta Kubera, Michael Maes

## Abstract

**Background:** There are strong associations between major depressive disorder (MDD), metabolic syndrome (MetS) and cardiovascular disorder, which may be explained by increased atherogenicity and the microimmuneoxysome (Maes et al., 1994; 2011). The present study was conducted to determine if MDD, the severity of depression, suicidal behaviors, and neuroticism are associated with increased pro-atherogenic versus anti-atherogenic indices (PRO/ANTI-AI) and a reverse cholesterol transport (RCT) index.

**Methods:** This study included 34 healthy controls, 33 participants with MetS, and MDD patients with (n=31) and without (n=35) MetS, and measured total (TC) and free (FC) cholesterol, high (HDLc) and low (LDLc) density lipoprotein cholesterol, triglycerides (TG), apolipoprotein (ApoA), ApoB, cholesterol esterification rate (CER) and a composite (based on HDLc, ApoA and CER), reflecting RCT.

**Results:** In the combined MDD + MetS study group, no associations between MDD and lipids were detected. After the exclusion of all MetS participants, MDD is strongly associated with (a) increased FC, TG, ApoB, Castelli risk index 1, ApoB/ApoA, and (b) decreased HDLc, ApoA and the RCT index. In participants without MetS, there are significant associations between severity of depression, suicidal behaviors, and neuroticism and ApoB/ApoA, Castelli risk, and RCT indices.

**Conclusions:** Studies linking lipids to depressive subtypes can only be interpreted after MetS patients are excluded. The depression phenome, suicidal behaviors, and neuroticism are associated with a lowered RCT and increased atherogenicity, which are likely involved in the microimmuneoxidative pathophysiology of MDD. The RCT is a new drug target to treat and prevent MDD, neuroticism, and suicidal behaviors.

## Introduction

There are strong associations between major depressive disorder (MDD), metabolic syndrome (MetS) and cardiovascular disorders (CVD) which in part may be explained by increased atherogenicity in MDD (de Melo et al., 2017; Maes et al., 1994; Maes et al., 1997; Morelli et al., 2021; Nunes et al., 2015). MetS is a collection of alterations in lipids, atherogenicity and insulin resistance that increase the risk for hypertension, CVD, type 2 diabetes mellitus (T2DM), and diabesity (late-life diabetes associated with obesity) (Lakka et al., 2002). Moreover, affective disorders, including MDD, are highly associated with obesity, dyslipidemia, and insulin resistance (Benton et al., 2007; Kupfer, 2005; Leboyer et al., 2012; Murphy et al., 1987) and may predispose, even at a young age, to MetS and accelerated atherosclerosis and CVD (Goldstein et al., 2015). Recent meta-analyses indicate that the prevalence of MetS is significantly elevated among those with MDD (Vancampfort et al., 2015). Individuals with MetS are more likely than those without to exhibit depressive symptoms (Capuron et al., 2008) and MetS comorbidity in mood disorders is associated with a more complex affective presentation, a lower likelihood of recovery, and an increase in the frequency of episodes and suicide attempts (Fagiolini et al., 2005; Fries et al., 2012; Grande et al., 2012; Thomas et al., 2008).

Both MDD and MetS are associated with elevated atherogenicity, as measured by an elevated Castelli risk index 1, which is the ratio of total cholesterol (TC) to high density lipoprotein cholesterol (HDLc), or an elevated atherogenic index of plasma (AIP), which measures the ratio of triglycerides (TG) to HDLc (Bortolasci et al., 2015; de Melo et al., 2017; Maes et al., 1994; Maes et al., 1997; Morelli et al., 2021; Nunes et al., 2015). Reduced blood HDLc levels, a lower cholesterol esterification rate (CER), and dysfunctions in reverse cholesterol transport (RCT) are key phenomena in MDD that partly explain elevated atherogenicity indices (Maes et al., 1994; Maes et al., 1997). HDLc prevents platelet aggregation, possesses antioxidant and anti-inflammatory characteristics, and protects mitochondria from reactive oxygen species (ROS) (Morris et al., 2021). Apoprotein A1 (ApoA1) and paraoxonase 1 (PON1) are the most essential proteins and enzymes that mediate the protective and anti-oxidant properties of HDL (Brites et al., 2017; Ferretti et al., 2010; Moreira et al., 2019a; 2019b; Morris et al., 2021). Cholesterol esterification is performed by lecithin:cholesterol acyl transferase (LCAT, EC 2.3.1.43), which esterifies cholesterol in plasma and prevents the inflow of free cholesterol (FC) into peripheral cells, hence promoting HDL maturation (Cooke et al., 2018; Morris et al., 2021). ApoA1 is produced by the liver and intestines, accumulates cholesterol, and activates LCAT, hence boosting the production and maturation of HDL (Ji et al., 2012; Kunnen and Van Eck, 2012; Zhou et al., 2015). According to Sadeghi et al., depression is accompanied by lowered ApoA levels (Sadeghi et al., 2011).

In addition, MDD may be associated with an increase in proatherogenic lipids and lipoproteins, such as low-density lipoprotein cholesterol (LDLc), TG, and ApoB (de Melo et al., 2017; Maes et al., 1994; Maes et al., 1997; Morelli et al., 2021; Nunes et al., 2015; Sadeghi et al., 2011). ApoB is a more inclusive indicator of all atherogenic lipoproteins, including LDL, very low density lipoprotein, and intermediate-density lipoprotein, due to the fact that each of these lipoproteins carries only one ApoB per particle (Attia, 2019; Sniderman, 2022). ApoB may better predict atherosclerosis, myocardial infarction, and cardiovascular disease than non-HDLc (Sniderman, 2022; Sniderman et al., 2019). There is currently evidence that tests of TG, TC, LDLc, and ApoB are required for accurate estimation of lipoprotein status (Sniderman et al., 2018) and that an elevated ApoB/ApoA ratio may be a superior atherogenicity index than LDLc, non-HDLc, and lipid ratios (Walldius and Jungner, 2006).

Already in the 1980s, there were indications that therapeutic trials aimed to lower serum cholesterol in coronary heart disease patients were counterbalanced by an increase in suicides and that depression is three times as prevalent among males aged 70 and older with low plasma cholesterol levels (Maes et al., 1994; 1997). In addition, other authors established that low cholesterol levels, especially in men, are linked to an increasing risk of medically serious suicide attempts (Sullivan et al., 1994). Nevertheless, Maes et al. (Maes et al., 1997) discovered that low HDLc, and not total cholesterol or LDLc, is connected with suicidal tendencies. Recent meta-analyses indicate that reduced antioxidant defenses, including HDLc levels, relate to current suicidal ideation and attempts (Vasupanrajit et al., 2021; Vasupanrajit et al., 2022).

Reportedly, Korean women with low HDLc are more likely to suffer from neuroticism (Roh et al., 2014), a personality trait characterized by increased sadness, anxiety, anger, worry, emotional instability, irritability, sensitivity to criticism, vulnerability to environmental stressors, and feelings of inadequacy (Goldberg, 1993; Lamb et al., 2002; Costa and McCrae, 1992; Watson et al., 1994). Intriguingly, statin therapy may marginally reduce the risk of neuroticism with an odds ratio of 0.90 (95% confidence interval of 0.83 to 0.97) (Alghamdi et al., 2018). We found that neuroticism (a trait), the phenome of depression (a state marker of the severity of MDD during the last week), and suicidal ideation and attempts (SI and SA) (state markers during the last month) are manifestations of the same latent vector (Jirakran et al., 2023). Therefore, the latter authors considered neuroticism to be a subclinical phenotype of MDD and that neuroticism, suicidal behaviors (SB), and depression should share biological pathways. However, it is unknown if suicidal behaviors and neuroticism are connected with RCT, decreased anti-atherogenicity and increased pro-atherogenicity levels, including the ApoB/A ratio.

Consequently, the present study was conducted to determine if MDD, the severity of depression, current suicidal ideation (SI), attempts (SA) and behaviors (SB), and neuroticism are associated with decreased RCT and anti-atherogenicity indices and elevated pro-atherogenicity indices. To control for the potential effects of MetS, we divided MDD patients and healthy controls into those with and without MetS and additionally controlled for the potential impact of age, sex, body mass index (BMI) and waist circumference.

## Methods and Participants

### Participants

This study included 133 participants, namely 66 major depressed patients and 67 age- and sex-matched controls, and of the 133 participants, 64 showed metabolic syndrome (MetS) and 69 did not have MetS. As such, we included 34 healthy controls, 35 MDD patients without MetS, 33 patients with MetS but no MDD, and 31 with MDD and MetS. We recruited male and female Thai-speaking participants between the ages of 18 and 65. Using the DSM-5 criteria, the patients were diagnosed with major depressive disorder (MDD) (American Psychiatric Association, 2013), and using the American Heart Association/National Heart, Lung, and Blood Institute 2009 Joint Scientific Statement (Alberti et al., 2009), we made the diagnosis of MetS. The Department of Psychiatry at King Chulalongkorn Memorial Hospital in Bangkok, Thailand, recruited outpatients from September 2021 to February 2022. The normal controls consisted of personnel, family or friends of personnel, and friends of MDD patients. Controls and patients were recruited from the same catchment area, namely Bangkok, Thailand.

Exclusion criteria for patients and controls are as follows: a) neurological, neuroinflammatory and neurodegenerative disorders, such as multiple sclerosis; stroke, Alzheimer and Parkinson disease, epilepsy, and brain tumors; b) major axis-1 diagnoses, such as schizophrenia, schizo-affective psychosis, autism, eating disorders, bipolar disorder, substance use disorders (SUDs), except tobacco use disorder (TUD); and psycho-organic disorders; c) axis-2 diagnoses, such as antisocial and borderline personality disorder; d) (auto)immune disorders, including inflammatory bowel disease, rheumatoid arthritis, chronic obstructive pulmonary disease, chronic kidney disease, systemic lupus erythematous, and psoriasis, e) abnormal kidney and liver function tests, f) lactating and pregnant women, g) lifetime use of immunosuppressants or immunomodulatory drugs, such as hydroxychloroquine, pomalidomide, and glucocorticoids, and h) use of therapeutic doses of omega-3 or antioxidant supplements. Moreover, healthy controls and MDD patients (without MetS) were excluded for using any lipid-lowering, anti-atherogenic, anti-diabetic, anti-hypertensive, and anti-obesity medications, including but not limited to statins, phenophybrate drugs, glipizide, metformin, amlodipine, enalapril, losartan, prazosin, and liraglutide. Nevertheless, patients with MetS and MDD+MetS were not excluded from using these medications. We also excluded normal controls if they suffered from lifetime or current DSM-5 dysthymia or DSM-IV anxiety disorders including generalized anxiety disorder (GAD), post-traumatic stress disorder (PTSD), obsessive-compulsive disorder (OCD), agoraphobia, social phobia, and panic disorder.

The research followed all relevant national and international privacy and ethical guidelines. The study was approved by the Institutional Review Board (#445/63) at Chulalongkorn University’s Medical Faculty in Bangkok, Thailand. Before taking part in the study, all subjects included were asked to voluntarily fill out a consent form that included detailed information about their participation.

### Clinical measurements

The clinical and sociodemographic data included in this study were collected using a semi-structured questionnaire administered to controls and patients. This interview includes socio-demographic data, family history (FHIS) of mood disorders and SUDs, psychotropic drugs used, and medical history. Using DSM-5 criteria (American Psychiatric Association, 2013) and the Mini International Neuropsychiatric Interview (M.I.N.I.) (Kittirattanapaiboon and Khamwongpin, 2005), MDD was diagnosed. In addition, the M.I.N.I. was used to make the lifetime and current diagnoses of dysthymia, panic disorder, GAD, social phobia, agoraphobia, OCD, PTSD and SUD. The 2009 Joint Scientific Statement of the American Heart Association and the National Heart, Lung, and Blood Institute defined Metabolic Syndrome (MetS) as the presence of three or more of the following components: (1) waist circumference ≥ 90 cm for men, ≥ 80 cm for women, or BMI ≥ 25 kg/m2; (2) high triglyceride level: ≥ 150 mg/dL; (3) low HDL cholesterol level: < 40 mg/dL for men, < 50 mg/dL for women; (4) high blood pressure: ≥ 130 mm Hg systolic blood pressure, ≥ 85 mm Hg diastolic blood pressure or treatment with antihypertensive medication; and (5) high fasting blood glucose ≥ 100 mg/dL or having a diabetes diagnosis. The formula for calculating the body mass index (BMI) is weight (kg) divided by height (m) squared.

The Hamilton Rating Scale for Depression (HAM-D, (Hamilton, 1960)) and the Beck Depression Inventory II (BDI-II) (Beck et al., 1996) were used to measure the severity of depression. Using the State-Trait Anxiety Inventory (STAI) state version (Spielberger et al., 1983) translated into Thai by Sompot et al. (Iamsupasit and Phumivuthisarn, 2005), the severity of anxiety was measured. As explained previously, we used principal component analysis (PCA) as a feature reduction method and extracted the first PC as an index of overall affective symptom severity (BDI, HAMD and STAI), dubbed phenome 1 (Jirakran et al., 2023; Maes et al., 2022a; Maes et al., 2022b). As explained, the first PC extracted from a data set is accepted as a validated construct only when: the factorability is adequate as assessed with the Kaiser-Meyer-Olkin test (satisfactory when > 0.5 an adequate when > 0.7), the Bartlett’s sphericity test is significant, the anti-image correlation matrix is adequate, the variance explained by the first PC is > 50%, and all loadings on the first PC are > 0.65. We utilised the Columbia Suicide Severity Rating Scale (C-SSRS) (Posner et al., 2011) to assess the severity of current SI and SA. The C-SSRS evaluates the severity of SI and SA, as well as self-injurious conduct without suicidal intent. As explained previously, we computed three indices of current suicidal behaviors, namely: a) the first PC extracted from C-SSRS items denoting current (last month) SI and frequency of SI; b) the first PC extracted from C-SSRS items denoting SA and frequency of SA; and c) a z composite score of SI + SA, dubbed suicidal behaviors (SB). We also constructed a second phenome score by extracting the first PC from BDI, HAMD, STAI and SB values, dubbed phenome 2.

The Big Five Inventory (BFI) (John and Srivastava, 1999) in a Thai translation (Luangsurong, 2016) was used to evaluate the five key personality traits of neuroticism, openness to experience, conscientiousness, extraversion, and agreeableness. In this study, we utilised the raw scores of the neuroticism dimension as only this domain is strongly associated with MDD (Jirakran et al., 2023). Tobacco use disorder (TUD) was diagnosed according to DSM-5 criteria.

#### Assays

Blood for the analysis of nitro-oxidative stress, immune and metabolic indicators was drawn at 8:00 a.m. following an overnight fast and within 48 h after admission into hospital. Aliquots of serum were kept at 80 °C until thawed for analysis. Using the Alinity C (Abbott Laboratories, USA; Otawara-Shi, Tochigi-Ken, Japan) with enzymatic (total cholesterol), accelerator selective detergent (HDL cholesterol), and glycerol phosphate oxidase (triglyceride) techniques, total and HDLc and TG were measured. The coefficients of variation for TC, HDLc, and TG were 2.3%, 2.6%, and 2.3%, respectively. The free cholesterol (FC) content was detected using the Free Cholesterol Colorimetric Assay Kit (Elabscience, cat number: E-BC-K004-M). All samples and reagents were performed on a 96-well plate with a plastic sealer and brought to room temperature before use. First, 5 μl of sterile distilled water was used as the blank well. 5 μl of the standard solution (reagent 2) was used as the standard well. Both reagents were performed in duplicate. 5 μl of serum was added to each well. Second, 250 μl of reagent 1 was then added to each well to avoid making an air bubble. Mix thoroughly, the plate was incubated at 37 °C for 10 minutes. The absorbance was measured at 510 nm using a microplate reader (Varioskan Flash Multimode, Thermo). The FC content was calculated according to the manufacturer’s instructions (Elabscience) and following the formula as 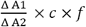 (mmol/L), with Δ A1: ODsample – ODblank, Δ A2: ODstandard – ODblank, c: concentration of the standard, f: dilution factor of sample before tested. Apo A1 and Apo B were measured from serum which has been centrifuged at 1500 g for 10 minutes. The serum was analyzed with an immunoturbidimetric assay technique via Roche Cobas 6000 with c501 module (Roche, Rotkreuz, Switzerland). The intra-inter assay coefficients of variation (CV) were 1.75-2.30% for Apo A1 and 2.64-4.51% for Apo B. Direct LDLc was measured using serum and the Liquid Selective Detergent method on the Alinity C machine by Abbot Laboratories, USA (manufactured in Otawara-shi, Tochigi-ken, Japan). The inter-assay CV for direct LDLc was 4.5%. We computed the ApoB/ApoA ratio and the Castelli risk index 1 as total cholesterol / HDLc. The esterified cholesterol ratio (ECR) was computed as: 1 – (free cholesterol / total cholesterol) x 100. This ratio estimates the cholesterol esterification rate (CER) (Maes et al., 1994). The cholesteryl ester (CE) levels were estimated by subtracting free cholesterol from the total cholesterol levels. We computed a new pro-atherogenicity index as: z transformation of ApoB (z ApoB) + z TG + z LDLc + z FC (dubbed PRO_AI). We computed a new z unit-weighted composite score reflecting anti-atherogenic potential (ANTI_AI) or RCT as: z HDLc + z ApoA + z CER. The primary atherogenicity index used in this study is the ratio of z PRO_AI – z ANTI_AI, dubbed as PRO/ANTI_AI.

### Statistical analysis

Chi-square or Fisher’s Exact Probability Test was used to compare nominal variables across categories. Analysis of variance (ANOVA) was used to compare scale variables between controls and MDD patients. Pearson’s product moment coefficients were used to calculate correlations between two sets of scale variables. We did not use p-corrections to account for the false discovery rate (FDR) when analyzing multiple comparisons or correlations between clinical and biomarker data, because these data are strongly intercorrelated and are not independent in statistical analysis. Using multiple regression analysis (manual approach), the most significant biomarkers (included as explanatory variables) for predicting the clinical data (e.g., phenome, neuroticism, SI and SB, entered as dependent variables), were identified. In addition, the best predictors of the model were identified using a forward stepwise automated regression method with p-values of 0.05 to-enter and 0.1 to delete. In the final regression models, in addition to F statistics (and p values) and total variance (R^2^ or partial eta squared as effect size), we estimated the standardized β coefficients with t-statistics and exact p-value for each of the explanatory variables. The White and modified Breusch-Pagan tests were used to verify the presence of heteroskedasticity. Collinearity and multicollinearity were examined using tolerance (cut-off value 0.25), variance inflation factor (cut-off value > 4), and the condition index and variance proportions from the collinearity diagnostics table. Partial regression plots were computed to display the independent association between dependent and explanatory variables. All of the above tests were two-tailed, and an alpha value of 0.05 was deemed statistically significant. In this study we use IBM, SPSS windows version 28.

## Results

### Socio-demographic and clinical data in MDD and controls

**Table 1** shows that there were no significant differences between MDD and controls in age, sex ratio, marital status, education, income, blood pressure, BMI, MetS prevalence, waist circumference, previous COVID-1 infection, and use of drugs to treat diabetes and MetS. Normal controls and MDD patients without MetS were free of any medical drugs, including antihypertensive and antidiabetic drugs, etc. The unemployment rate, BDI, HAMD, STAI as well as phenome 1 and phenome 2, and the current SI, SA and SB scores were significantly higher in MDD than in controls. Part of the patients were taking psychotropic drugs, including sertraline (n=28), trazodone (n=16), fluoxetine (n=7), venlafaxine (n=13), paroxetine (n=2) and agomelatine (n=2) mirtazapine (n=8), benzodiazepines (n=36) and mood stabilizers (n=2).

**Table 1.** Demographic and clinical data of the major depression patients (MDD) and healthy controls (HC) included in the present study.

| Variables | HC (n=67) | MDD (n=66) | F/X<sup>2</sup>/FEPT | df | p |
| --- | --- | --- | --- | --- | --- |
| Sex (Male / Female) | 9/58 | 18/48 | 3.94 | 1 | 0.055 |
| Age (years) | 37.9 (9.2) | 36.9 (11.5) | 0.263 | 1/131 | 0.609 |
| Education (years) | 14.1 (3.4) | 14.8 (2.8) | 1.72 | 1/131 | 0.193 |
| Income (baht/month) | 21500 (7269) | 23770 (19203) | 0.817 | 1/131 | 0.368 |
| Single/married/widower/divorced | 35/26/3/3 | 36/22/4/4 | FHHET |  | 0.877 |
| Employed / unemployed | 66/1 | 47/19 | 19.388 | 1 | <0.001 |
| TUD (No/Yes) | 62/5 | 54/12 | 3.43 | 1 | 0.074 |
| BMI (kg/m<sup>2</sup>) | 27.4 (6.1) | 26.9 (5.5) | 0.266 | 1/130 | 0.607 |
| Waist Circumference (cm) | 88.8 (14.9) | 89.7 (15.0) | 0.142 | 1/130 | 0.707 |
| Systolic Blood Pressure (mmHg) | 127.9 (15.3) | 129.9 (17.8) | 0.475 | 1/131 | 0.492 |
| Diastolic Blood Pressure (mmHg) | 79.2 (11.3) | 77.9 (12.4) | 0.420 | 1/131 | 0.518 |
| Metabolic syndrome (MetS) (No/Yes) | 34/33 | 35/31 | 0.069 | 1 | 0.863 |
| Neuroticism | 18.3 (5.0) | 27.2 (6.4) | 80.332 | 1/131 | <0.001 |
| Total BDI | 5.6 (6.5) | 26.9 (13.8) | 129.518 | 1/131 | <0.001 |
| Total HAMD | 2.3 (3.0) | 18.1 (6.4) | 330.943 | 1/131 | <0.001 |
| Total STAI | 38.2 (7.8) | 51.9 (10.4) | 73.623 | 1/131 | <0.001 |
| PC HAMD + BDI + STAI | -0.758 (0.421) | 0.770 (0.810) | 187.443 | 1/131 | <0.001 |
| PC HAMD + BDI + STAI + Current SB | -0.754 (0.352) | 0.765 (0.850) | 182.045 | 1/131 | <0.001 |
| Current suicidal ideation (SI) | -0.489 (0.0) | 0.493 (1.248) | KWT | - | <0.001 |
| Current suicidal attempts (SA) | -0.245 (0.0) | 0.256 (1.390) | KWT | - | 0.004 |
| Current Suicidal behaviors (SI and SA) | -0.420 (0.0) | 0.426 (1.290) | KWT | - | <0.001 |
| Previous Covid-19 infection (No/Yes) | 62/5 | 62/4 | 0.104 | 1 | 1.000 |
| T2DM (No/Yes) | 61/6 | 61/5 | 0.083 | 1 | 1.000 |
| Dyslipidemia (No/Yes) | 58/9 | 55/11 | 0.272 | 1 | 0.635 |
| Drugs targeting MetS-related conditions (No/Yes) | 50/17 | 52/14 | 0.322 | 1 | 0.682 |

Results are shown as mean ± SD. F: results of analysis of variance; X<sup>2</sup>: analysis of contingency tables, FFHT: Fisher-Freeman-Halton Exact Test; KWT: Kruskal-Wallis test.
BMI: body mass index; TUD: Tobacco use disorder; BDI: Beck Depression Inventory; HAMD: Hamilton Depression Rating Scale; STAI: State and Trait Anxiety Inventory; PC: principal component; SB: suicidal behaviors, that is ideation (SI) and attempts (SA); T2DM: type 2 diabetes mellitus; PC HAMD + BDI + STAI: first PC score extracted from these three rating scales; PC HAMD + BDI + STAI + Current SB: first PC score extracted from these three rating scales and suicidal behaviors

### Lipid differences between MDD and controls

**Table 2** shows that there were no significant differences between MDD and controls in any of the lipid levels (HDLc, TC, FC, CE, LDLc, TG, and their ratios or composites (CER, ApoB/ApoA, Castelli risk 1 index, with or without covarying for age, sex, BMI and waist circumference. Nevertheless, after performing the same analyses in those without MetS showed completely different results. **Table 3** shows the results of univariate GLM analyses performed on subjects without MetS, thus comparing MDD without MetS with controls without MetS. We found that FC, TG, ApoB, Castlelli risk index 1, ApoB/ApoA PRO_AI, and PRO/ANTI_AI were significantly higher in MDD patients than in controls. HDLc, ApoA, and ANTI_AI were significantly lower in MDD patients than in controls. Covarying for the antidepressant drug state and benzodiazepines did not change any of these results and showed that MDD remained significant and that the drug state was non-significant even after FDR p-correction.

**Table 2.** Biomarker data of the major depressed patients (MDD) and healthy controls (HC) included in this study.

| Variables | HC (n=67) | MDD (n=66) | F | df | p |
| --- | --- | --- | --- | --- | --- |
| Total cholesterol (mg/dL) | 205.6 (4.9) | 202.9 (5.0) | 0.14 | 1/125 | 0.708 |
| Free cholesterol (mg/dL) | 65.4 (2.7) | 63.3 (2.8) | 0.28 | 1/125 | 0.596 |
| Cholesteryl esters (mg/dL) | 140.1 (4.1) | 139.6 (4.2) | 0.01 | 1/125 | 0.927 |
| Cholesterol esterification ratio (%) | 68.6 (1.1) | 68.2 (1.2) | 0.05 | 1/125 | 0.822 |
| HDLc (mg/dL) | 55.2 (1.5) | 53.7 (1.5) | 0.52 | 1/125 | 0.472 |
| Triglycerides (TG) (mg/dL) * | 180.2 (21.9) | 135.1 (22.4) | 0.00 | 1/125 | 0.960 |
| LDLc (mg/dL) | 128.2 (4.6) | 135.3 (4.7) | 1.16 | 1/125 | 0.284 |
| Apolipoprotein A (ApoA) (mg/dL) | 142.6 (3.3) | 135.3 (3.3) | 2.37 | 1/125 | 0.126 |
| ApoB (mg/dL) | 95.2 (3.1) | 98.6 (3.1) | 0.59 | 1/125 | 0.444 |
| ApoB/ApoA ratio * | 0.70 (0.03) | 0.75 (0.03) | 1.52 | 1/125 | 0.221 |
| Castelli risk index 1 | 4.02 (0.14) | 3.97 (0.14) | 0.07 | 1/125 | 0.791 |
| TG/HDLc ratio | 3.98 (0.59) | 2.71 (0.60) | 2.23 | 1/125 | 0.138 |

Results are shown as estimated marginal means (SEM) after univariate GLM analyses with age, sex, body mass index and waist circumference as covariates. \* Processed in Log-transformation; HDLc: high density lipoprotein cholesterol; LDLc: low density lipoprotein cholesterol

**Table 3.** Biomarker data of the major depression patients (MDD) and healthy controls (HC) without metabolic syndrome.

| Variables | HC (n=34) | MDD (n=35) | F | df | p |
| --- | --- | --- | --- | --- | --- |
| Total Cholesterol (mg/dL) | 190.0 (6.3) | 202.8 (6.3) | 2.02 | 1/62 | 0.160 |
| Free Cholesterol (FC) (mg/dL) | 51.7 (3.2) | 62.0 (3.2) | 5.00 | 1/62 | 0.029 |
| Cholesteryl esters (mg/dL) | 138.3 (5.6) | 140.8 (5.6) | 0.10 | 1/62 | 0.756 |
| Cholesterol esterification rate (CER) | 72.8 (1.5) | 69.2 (1.5) | 2.72 | 1/62 | 0.104 |
| HDLc (mg/dL) | 64.3 (2.4) | 56.8 (2.4) | 4.83 | 1/62 | 0.032 |
| Triglycerides (TG) (mg/dL) * | 84.1 (8.9) | 123.1 (8.9) | 9.71 | 1/62 | 0.003 |
| LDLc (mg/dL) | 118.2 (6.2) | 132.3 (6.2) | 2.52 | 1/62 | 0.118 |
| Apolipoprotein A (ApoA) (mg/mL) | 149.8 (5.0) | 135.2 (5.0) | 4.14 | 1/62 | 0.046 |
| ApoB (mg/mL) | 80.6 (3.6) | 91.5 (3.6) | 4.39 | 1/62 | 0.040 |
| ApoB/ApoA ratio * | 0.56 (0.03) | 0.70 (0.03) | 10.34 | 1/62 | 0.002 |
| Castelli risk index 1 | 3.05 (0.16) | 3.74 (0.16) | 9.59 | 1/62 | 0.003 |
| TG/HDLc | 1.37 (0.22) | 2.39 (0.22) | 10.75 | 1/62 | 0.002 |
| PRO_AI (z scores) | -0.308 (0.170) | 0.325 (0.170) | 6.73 | 1/62 | 0.012 |
| ANTI_AI (z scores) | 0.308 (0.165) | -0.286 (0.165) | 6.29 | 1/62 | 0.015 |
| PRO/ANTI_AI (z scores) | -0.392 (0.163) | 0.384 (0.163) | 11.00 | 1/62 | 0.002 |

Results are shown as estimated marginal means (SEM) after univariate GLM analyses with age, sex, body mass index and waist circumference as covariates; \* Processed in Log transformation; HDLc: high density lipoprotein cholesterol; LDLc: low density lipoprotein cholesterol; PRO\_AI: composite based on LDLc, TG, ApoB, and FC; ANTI\_AI: composite based on HDLc, ApoA and CER; PRO/ANTI\_AI: PRO\_AI versus ANTI\_AI ratio

Electronic Supplementary File (ESF), Table 1 shows the measurements in patients with MetS with MDD versus those with MetS without MDD. There were no significant differences in any of the lipid, composites, or ratios between MetS and MetS+MDD. ESF, Table 2 shows the differences in lipid profiles between controls and people with MetS, after excluding all patients with MDD. We found significantly higher levels of all atherogenic and lowered levels of all anti-atherogenic lipid (except CE, ApoA) variables in MetS versus controls.

ESF, Figure 1 shows the levels of FC in MDD and controls (results of an univariate GLM analysis), considering the significant interaction term between MDD and MetS (F=7.53, df=1/123, p=0.007). Comparisons of simple main effects showed that in patients with MetS, FC was lower in those with MDD, while there was a trend towards an opposite association in those without MetS (p=0.025). ESF, Figure 2 shows HDLc in MDD and controls and that the MDD x MetS interaction was significant (F=5.79, df=1/123, p=0.018). The results show that MDD patients without MetS have significantly lower HDLc values than controls (p=0.022), whereas MDD patients with MetS show a trend towards increased HDLc as compared with MetS without MDD. No such interaction patterns were established for TC, CER, TG, ApoA, ApoB and LDLc. **Figure 1** shows the ApoB/ApoA ratio in MDD and MetS and the significant MDD x MetS interaction (F=6.36, df=1/123, p=0.013). In those without MetS, the ratio was significantly higher in people with MDD as compared with controls (p=0.003) and, in fact, the ratio was not significantly different between MDD and MetS with or without MDD. ESF, Figure 3 shows the Castelli risk index 1 in MDD and MetS patients and a significant MDD X MetS interaction (F=11.23, df=1/123, p=0.001); in subjects without MetS, there was a significantly higher index in MDD as compared with controls (p=0.030), whereas in those with MetS there was a significant decrease in the ratio (p=0.014). ESF, Figure 4 shows the PRO_AI index in MDD and MetS and the significant MDD X MetS interaction (F=7.00, df=1/123, p=0.009); in subjects without MetS, there was a trend towards a higher PRO_AI index in MDD patients as compared with controls (p=0.064), whereas in those with MetS there was a trend towards a lower PRO_AI index in MDD patients (p=0.069). **Figure 2** shows the ANTI_AI or RCT index in MDD and MetS as well as the significant interaction pattern between both diagnostic groups (F=5.95, df=1/123, p=0.016); in subjects without MetS, there was a significantly lower index in patients with MDD as compared with controls (p=0.011), whereas in those with MetS there was a trend towards an increase in MDD patients (p=0.381). ESF, Figure 5 shows the PRO/ANTI_AI in MDD and MetS and the significant interaction pattern (F=11.18, df=1/123, p=0.001); in people without MetS, this ratio was significantly higher in MDD patients as compared with controls (p=0.004), whereas in people with MetS, there was a trend toward a lowered index in MDD patients (p=0.078).

**Figure 1.**
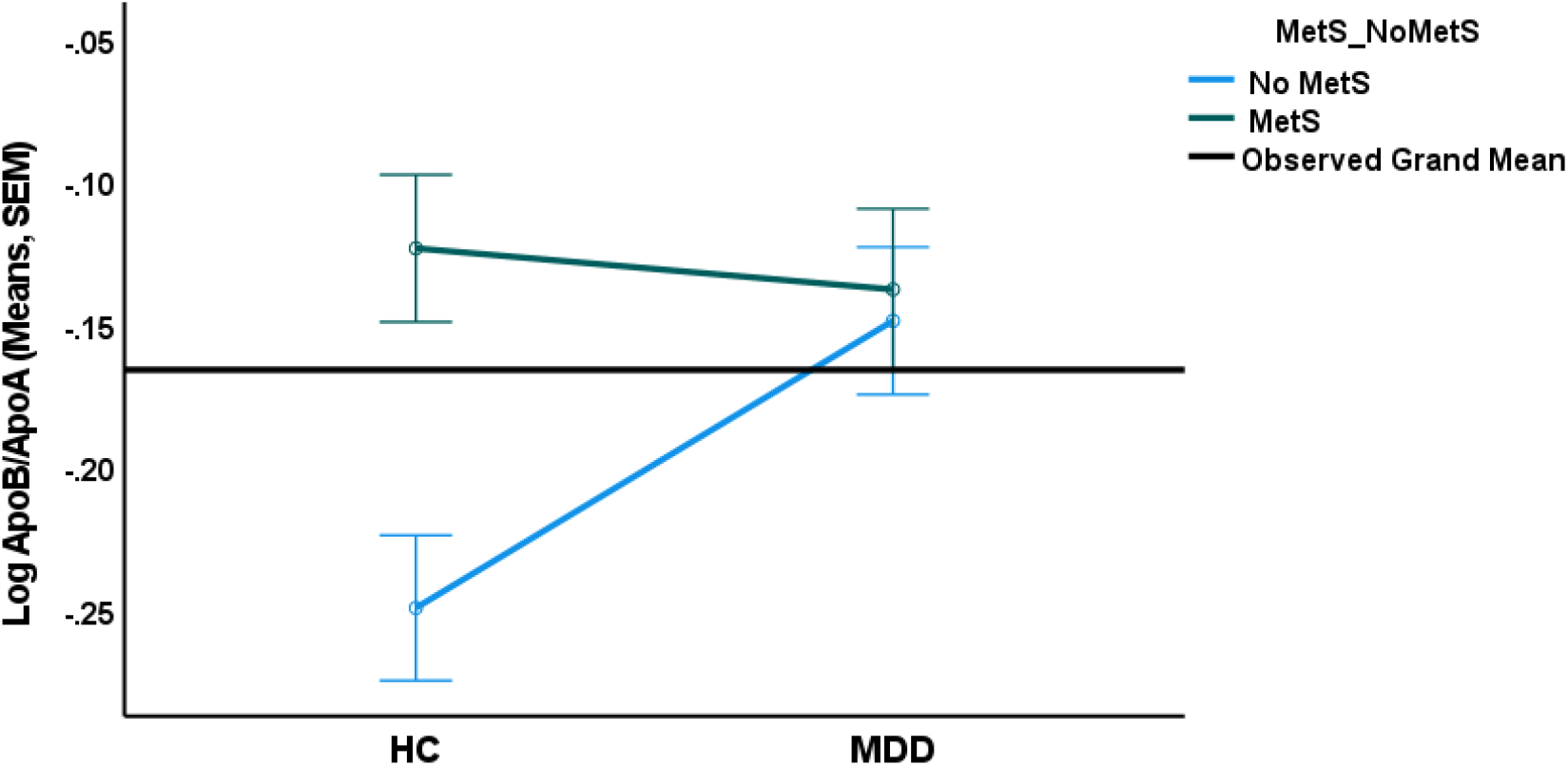
Interaction pattern between major depression (MDD versus healthy controls, HC) and metabolic syndrome (MetS). Shown are the estimated marginal mean values of the apolipoprotein (Apo)B/ApoA ratio (in logarithmic transformation) after covarying for age, sex, body mass index and waist circumference

**Figure 2.**
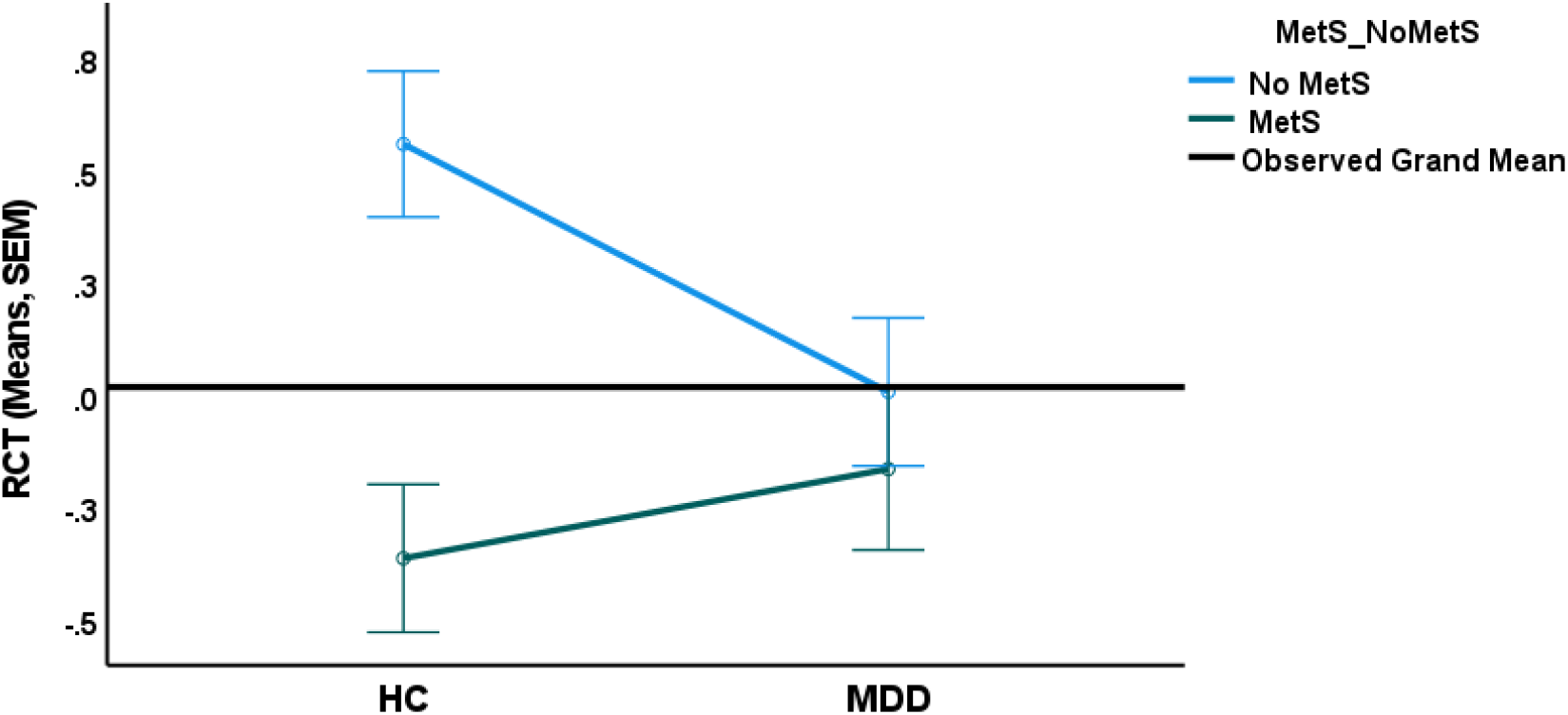
Interaction pattern between major depression (MDD versus healthy controls, HC) and metabolic syndrome (MetS). Shown are the estimated marginal mean values of the reverse cholesterol transport (RCT) index after covarying for age, sex, body mass index and waist circumference

**Figure 3.**
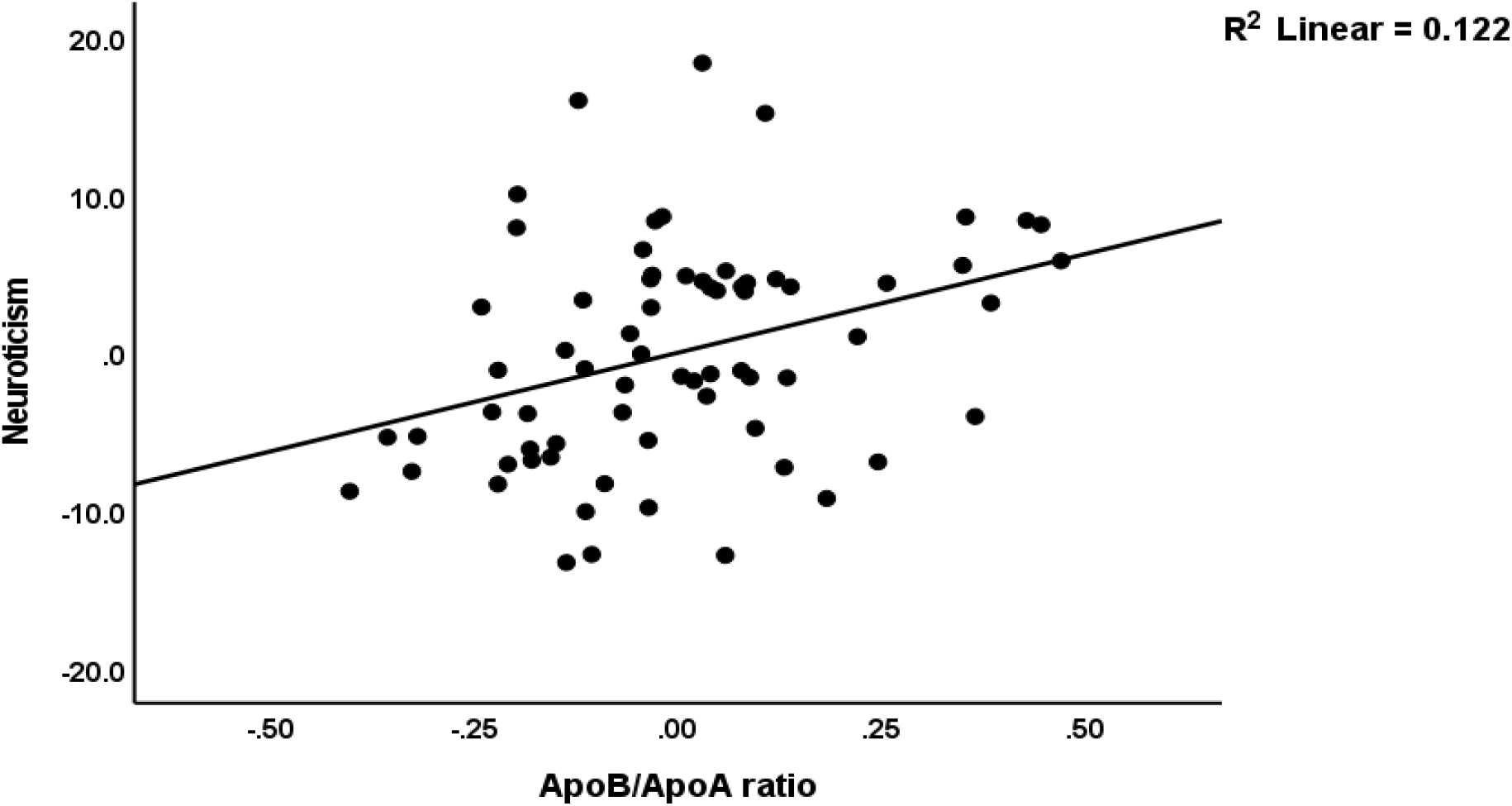
Partial regression of the first principal component extracted from depression, anxiety and suicidal behaviors scores and the apolipoprotein (Apo)B/ApoA ratio

**Figure 4.**
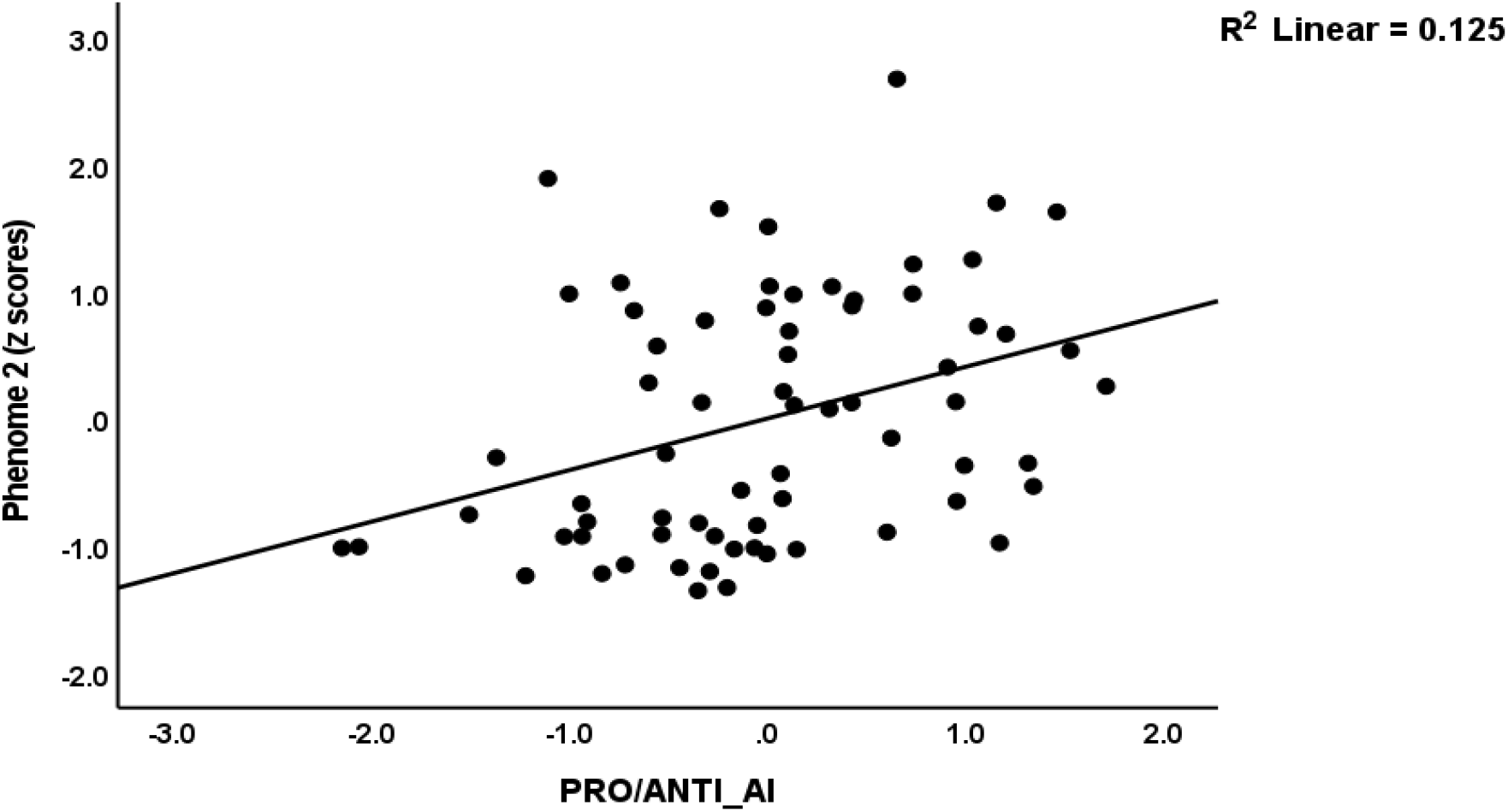
Partial regression of the first principal component extracted from depression, anxiety and suicidal behaviors scores and the PRO_AI versus ANTI_AI ratio, with PRO_AI: composite based on low density lipoprotein cholesterol, triglycerides, apolipoprotein (Apo)B, and free cholesterol (FC); ANTI_AI: composite based on high density lipoprotein cholesterol, ApoA and cholesterol esterification rate

### Intercorrelation matrix

**Table 4** shows the intercorrelation matrix between the important lipid composites or ratios and clinical variables. We found that ANTI_AI and PRO/ANTI_AI were significantly associated with neuroticism, HAMD, STAI, phenome 1, SI, SB, whereas PRO_AI was only associated with SI (at p=0.032). The ApoB/ApoA index was significantly correlated with all clinical variables.

**Table 4.** Correlation matrix between atherogenic indices and clinical depression data.

| Variables | PRO_AI | ANTI_AI | PRO_ANTI ratio | ApoB/ApoA |
| --- | --- | --- | --- | --- |
| Neuroticism | 0.177 (0.146) | -0.264 (0.029) | 0.330 (0.006) | 0.347 (0.004) |
| Total BDI | 0.091 (0.457) | -0.213 (0.078) | 0.267 (0.027) | 0.267 (0.027) |
| Total HAMD | 0.182 (0.134) | -0.285 (0.018) | 0.354 (0.003) | 0.365 (0.002) |
| Total STAI | 0.102 (0.403) | -0.356 (0.003) | 0.370 (0.002) | 0.363 (0.002) |
| PC HAMD + STAI + BDI | 0.134 (0.274) | -0.303 (0.011) | 0.352 (0.003) | 0.353 (0.003) |
| Current SI | 0.259 (0.032) | -0.249 (0.039) | 0.312 (0.009) | 0.251 (0.038) |
| Current SA | 0.080 (0.514) | -0.292 (0.015) | 0.260 (0.031) | 0.194 (0.111) |
| Current SB (SI+SA) | 0.193 (0.113) | -0.297 (0.013) | 0.318 (0.008) | 0.247 (0.041) |
| PC HAMD + BDI + STAI + CurrentSB | 0.175 (0.150) | -0.316 (0.008) | 0.373 (0.002) | 0.359 (0.002) |

Results of Pearson correlation calculations. BDI: Beck Depression Inventory; HAMD: Hamilton Depression Rating Scale; STAI: State and Trait Anxiety Inventory; PC: principal component; SB: suicidal behaviors, that is suicidal ideation (SI) and attempts (SA); PC HAMD + STAI + BDI: first PC score extracted from these three rating scales; PC HAMD + BDI + STAI + CurrentSB: first PC score extracted from these three rating scales and suicidal behaviors
PRO\_AI: composite based on low density lipoprotein cholesterol, triglycerides, apolipoprotein (Apo)B, and free cholesterol; ANTI\_AI: composite based on high density lipoprotein cholesterol, ApoA and cholesterol esterification rate; PRO/ANTI\_AI: PRO\_AI versus ANTI\_AI ratio.

### Results of multiple regression analysis

**Table 5** shows the results of different multiple regression analyses performed in subjects without MetS with clinical ratings as dependent variables and lipids as explanatory variables, while allowing for the effects of age, sex, BMI an waist circumference. Regression #1 shows that 20.2% of the variance in current SI is explained by TG (positively) and age (inversely). Regression #2 shows that 32.2% of the variance in phenome 1 is explained by the regression on TG (positively) and ApoA and age (both inversely). 35.0% of the variance in phenome 2 was explained by TG, ApoA and age. We found that 21.1% of the variance in neuroticism was explained by the regression on ApoB/ApoA ratio (positively) and age (inversely). **Figure 3** shows the partial regression of neuroticism on the ApoB/ApoA ratio. We have rerun the same analyses with the three composites (PRO_AI, ANTI_AI and PRO/ANTI_AI) as explanatory variables and with age, sex, BMI and waist circumference as explanatory variables. Table 5, regression #5 shows that 16.5% of the current SI variance was explained by PRO/ANTI_AI (positively) and age (inversely). Up to 23.4% and 25.7% of the variances in phenome 1 and phenome 2, respectively, were explained by PRO/ANTI_AI and age. **Figure 4** shows the partial regression of phenome 2 on PRO/ANTI_AI. The same two input variables also explained 18.8% of the variance in neuroticism. Consequently, we performed the same regression in subjects with MetS, but no significant associations between the clinical data and any of the lipid data, ratios or composites could be detected.

**Table 5.**
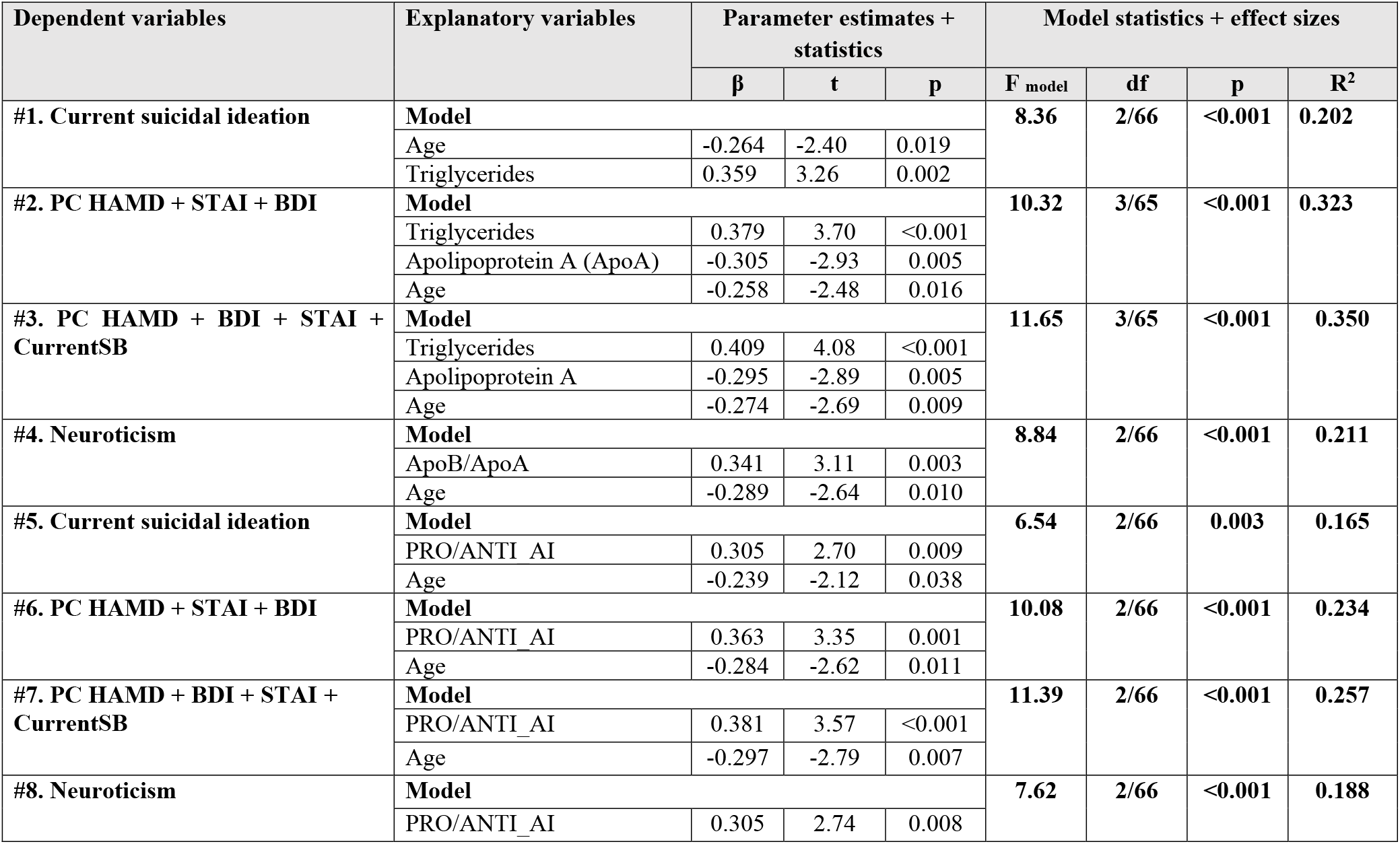
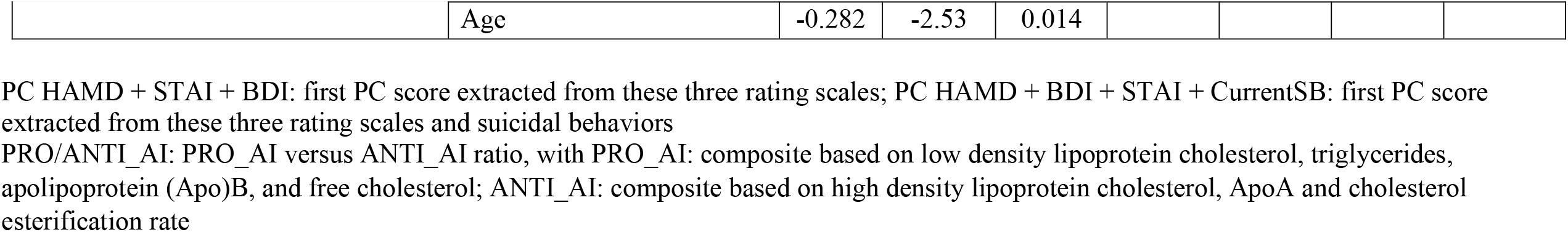
Results of multiple regression analyses with clinical data as dependent variables and biomarkers as explanatory variables

## Discussion

### Lipid profiles of MDD, suicidal behaviors and neuroticism

The most important finding of this study is that MDD (after exclusion of cases with MetS) is strongly associated with (a) increased pro-atherogenic indices, including the Castelli risk index 1, ApoB/ApoA ratio and PRO/ANTI_AI, as well as increased levels of FC, TG and ApoB; and (b) decreased levels of anti-atherogenic lipids, including HDLc and ApoA and the RCT index. Maes et al. (Maes et al., 1994) reported the first evidence of increased atherogenicity in relation to major depressive disorder by observing a lower HDLc/TC ratio (the inverse of the Castelli risk index 1) in MDD. Other researchers have reported increased atherogenic indices in MDD (Bortolasci et al., 2015; Morelli et al., 2021; Nunes et al., 2015). Maes et al. (Maes et al., 2011) reviewed that increased atherogenicity coupled with oxidative and inflammatory mechanisms underpin the co-occurrence of MDD, atherosclerosis, and CVD (Maes et al., 2011). In addition, elevated atherogenicity mediates the effects of other medical conditions, such as T2DM and unstable angina, on depressive symptoms due to these medical conditions (Al-Hakeim et al., 2022; Mousa et al., 2022).

In addition to elevated TG, the current study demonstrates that elevated levels of FC are a crucial component of MDD, whereas there are no differences in TC, LDL, or CE between MDD patients and controls. To our knowledge, no other FC and CE-related papers have been published in MDD. As Maes et al. (Maes et al., 1994), the current study established that low RCT is a key feature of MDD, as measured by low HDLc, ApoA and CER. In the present study, there was a trend towards lower CER in MDD, although previously we detected a significantly lowered CER in MDD and family members of depressed patients, indicating diminished LCAT activity (Maes et al., 1994). However, CE and the CER are probably not the best indicators of LCAT activity, as CE’s fate is dependent on the CE contents of LDL and VLDL, the activity of cholesteryl ester transfer protein (CETP), and the liver’s uptake of HDL and CE (Gauthier et al., 2005). Nonetheless, our findings that FC is elevated and HDLc is decreased (LCAT promotes HDLc maturation; see Introduction) further underscore that LCAT activity may be diminished (Sniderman et al., 2019) in MDD (Maes et al., 1994).

Clinically, the ApoB/ApoA and Castelli risk ratios, as well as the newly developed composite score comprising all important lipid biomarkers, namely PRO/ANTI_AI were the best predictors of MDD. When comparing the important effect size of the ApoB/ApoA ratio versus its single indicators, AopA or ApoB, which are only marginally different between MDD patients and controls, the importance of those ratios for MDD becomes clear. From a practical standpoint, the ApoB/ApoA ratio and Castelli index are the most useful because these ratios are easy to measure, whereas the z composite scores may be more difficult to compute in clinical practice.

In the current study, we also determined that elevated atherogenicity (including TG, ApoB/ApoA ratio, PRO/ANTI_AI), and decreased RCT are significantly associated with current suicidal ideation and attempts. These results extend those of Maes et al. (Maes et al., 1997), who reported a significant association between HDLc and suicidal ideation. A recent meta-analysis and systematic review demonstrated that low HDLc contributes to the significant association between neuro-immune-oxidative stress and current suicidal ideation (Vasupanrajit et al., 2022). In addition, we discovered that neuroticism was inversely associated with RCT, and positively with the ApoB/ApoA ratio, and PRO/ANTI-AI. These findings suggest that neuroticism, MDD, and suicidal behaviors may share a common pathophysiology, and that neuroticism is a subclinical phenotype of MDD (Jirakran et al., 2023).

### Issues when measuring lipids in MDD

A key finding of the current study is that when MDD patients and controls are combined with MetS patients in one study sample, the actual associations between MDD and lipids become completely muddled. Covarying for MetS cannot eliminate the confounding effects of MetS, although a factorial ANCOVA approach with MetS as second factor, age, sex, and BMI as covariates is adequate, and additionally may detect the interaction patterns MDD x MetS. Thus, if too many cases of MetS are included in the study groups of patients and controls, the actual significance will disappear, or a significant effect may appear when more people with MetS are included in MDD versus control groups. The message is that when comparing the lipid profile in MDD versus controls, all subjects with MetS should be excluded, and the results should be adjusted for age, sex, BMI, and waist circumference.

We are currently undertaking a meta-analysis on the lipid profiles of MDD and have discovered that only few studies have excluded people with MetS, whereas some have removed subjects with a high BMI. For example, only 16.9% of studies reporting atherogenic lipid biomarkers mentioned whether individuals with MetS were included, and those that did not exclude MetS, did not account for MetS (Jirikran et al., to be submitted, February 2023). In addition, around 27.2% of the studies did not control for BMI, and many studies (almost 43%) did not report on the timing of blood collections, and of those that did, the majority of participants were often not fasting (Jirakran et al., 2023). Tests such as LDLc, HDLc, and TG require fasting, best 12 hours (ApoB Test, 2021). Moreover, when nonfasting assays of TC, LDLc, non-HDLc, and ApoB are used, the predictive value for incident CVD is limited (Mora et al., 2008).

### Lipids and the pathophysiology of MDD

In the current study, the key outcome variables were z composite scores used to generate pro- and anti-atherogenic indices. As previously discussed, the pro-atherogenic indicators utilized in this study (TC, FC, LDLc, TG, and ApoB) are interrelated and cannot be considered independent variables in statistical analysis (Sniderman et al., 2019). ApoB is likely one of the most accurate predictors of incident cardiovascular disease, although one must also examine the interplay between ApoB and TG, LDLc, and TC (Sniderman et al., 2019). Therefore, the most adequate method is to compute z unit-weighted composite scores that account for the effects of all important pro-atherogenic players.

FC contributes to atherosclerosis and the formation of atheroma because it can diffuse directly into the arterial wall, induce cytotoxicity, suppress the formation of membrane domains, may accumulate and crystallize in cells and macrophages, promote the oxidation of cholesterol to oxysterols, some of which may inhibit FC efflux, and induce apoptotic pathways and cell death (Tabas, 2002). LDLc is an atherogenic lipoprotein that undergoes oxidative alteration to become oxidized LDL (oxLDL), which initiates inflammatory and autoimmune responses that drive atherogenic responses (Maes et al., 2011). In addition to reflecting increases in LDL and VLDL, elevated levels of ApoB may directly induce atherosclerosis (Behbodikhah et al., 2021; Sniderman et al., 2019). Elevated TG levels are independent risk factors for CVD by reducing HDLc, raising LDLc, activating plasminogen activators, and thrombogenic factors (McBride, 2008). In fact, patients with elevated TG and LDLc levels and low HDLc levels are more likely to develop CVD.

Similarly, we computed a z unit-weighted composite score reflecting anti-atherogenic effects (HDLc and ApoA) and included the CER (indicating LCAT activity), and, therefore, this composite reflects RCT. ApoA inhibits apoptosis and platelet activation, induces vasodilation, and promotes innate immunity (Georgila et al., 2019). Not only are RCT, ApoA, and HDLc anti-atherogenic, but they have anti-inflammatory and antioxidant properties (Madison et al., 2021; Moreira et al., 2019a; 2019b; Murphy et al., 2011).

As a result, the current study’s findings are extremely significant in terms of the microimmuneoxysome pathophysiology of MDD, which includes the interplay between immune ad oxidative pathways, and the gut microbiome with increased gut permeability and translocation of lipopolysaccharides (LPS) of Gram-negative gut commensals (Maes, 1995; Maes et al., 1990; Maes and Carvalho, 2018; Maes et al., 2008; Maes et al., 2023). The ApoA-HDLc complex has antiviral activity and may neutralize LPS, thereby attenuating the Toll-Like Receptor 4 complex and inflammatory responses (Georgila et al., 2019). Decreased LCAT activation may further reduce HDLc’s LPS-neutralizing effects (Petropoulou et al., 2015). Furthermore, low ApoA-HDLc levels may contribute to chronic inflammation, whereas ApoA inhibits dendritic cell maturation and differentiation, inhibits the NLRP3 inflammasome, pro-inflammatory T helper-(Th)1 and Th17 profiles, interleukin (IL)-1β, IL-12, and interferon-gamma, and increases IL-10, an anti-inflammatory cytokine (Georgila et al., 2019). As a result, the decreased RCT and lower ApoA and HDLc levels may contribute to the activated M1, Th1, Th2, and Th17 pathways, as well as the activated oxidative pathways, in MDD (Maes et al., 2018; Maes and Carvalho, 2018).

Nevertheless, activation of immune-inflammatory and oxidative pathways has detrimental effects on the HDLc-PON1 complex and on ApoA leading to lowered activity of the protective capacities and even transforming this anti-inflammatory complex into a pro-inflammatory complex (Georgila et al., 2019; Maes et al., 2018; Moreira et al., 2019b; Morris et al., 2021; Van Lenten et al., 1995). Probably, the disorders in RCT and atherogenicity are driven by (genetic) attenuation of PON1 activity (Siroka et al., 2020), which is a key characteristic of MDD that is determined (in part) by PON1 Q192R genetic variants (Moreira et al., 2019a; 2019b). Thus, the latter may drive dysfunctions in the HDL-PON1-ApoA-LCAT complex, leading to lowered RCT, which in turn may cause increased oxidative and inflammatory responses, which, in turn, affect the HDL-PON1-ApoA-LCAT complex (Maes et al., 2018). Thus, reciprocal interrelations between the HDL-PON1-ApoA-LCAT complex and the microimmuneoxysome are probably key components of MDD.

### Lipids, MetS and MDD

The study’s third major finding was that MetS was associated with changes in all lipid biomarkers except CE and ApoA. The most important MetS biomarkers in our study were the increased PRO/ANTI_AI, Castelli risk index 1, and TG, as well as lower HDLc levels. These findings are consistent with previous reports that MetS is associated with increased TC, TG, Castelli risk index 1, and decreased HDLc (Haile et al., 2021; Kawamoto et al., 2011; Paredes et al., 2019). Other studies have found that MetS is associated with increased ApoB and decreased RCT (Lim et al., 2015; Sviridov et al., 2008). This significant overlap in lipid profiles between MetS and MDD not only explains the comorbidity between the two conditions, but also why all subjects with MetS must be excluded when assessing the lipid profile of MDD patients.

Other findings of the current study, however, show an even more important reason to exclude subjects with MetS, namely the presence of significant interaction patterns between MDD and MetS, wherein subjects without MetS have a significant increase in ApoB/ApoA, Castelli risk index1, PRO_AI, PRO/ANTI_AI in MDD, and a significant decrease in RCT and HDLc as compared to controls, whereas those with MetS show often the opposite association. This suggests that people with MDD + MetS have a less severe lipid profile than people with MetS alone. As a result, MDD appears to reduce the abnormal lipid profile that has been established in MetS. Nonetheless, these findings are based on an exploratory observation, and not on *a priori* hypothesis testing. MDD combined with MetS results in increased nitro-oxidative stress with lipid peroxidation and chlorinative stress, both of which are associated with increased atherogenicity (Morelli et al., 2021). As a result, future research should look into the interactions between MDD and MetS and examine possible moderating effects on atherogenicity, inflammation and nitro-oxidative stress.

## Conclusions

There are no correlations between MDD and lipids in the overall MDD + MetS study population. MDD is highly linked with (a) increased FC, TG, ApoB, Castelli risk index 1, ApoB/ApoA, and PRO/ANTI_AI and (b) decreased HDLc, ApoA, and the RCT index after excluding all patients with MetS. In participants without MetS, the severity of depression, suicidal behaviors, and neuroticism are significantly correlated with RCT, indicating that increased atherogenicity is, at least in part, driven by lowered RCT. PON1 Q192R genetic variants may drive the aberrations in the HDL-PON1-ApoA-LCAT complex, which may cause increased oxidative and inflammatory responses, which affect the HDL-PON1-ApoA-LCAT complex. Consequently, reciprocal interrelationships between the HDL-PON1-ApoA-LCAT complex and the microimmuneoxysome are likely essential components of the phenome of depression, neuroticism and suicidal behaviors. The HDL-PON1-ApoA-LCAT complex, RCT and increased atherogenicity in conjunction with the microimmuneoxysome are new drug targets to treat and prevent MDD, neuroticism, and suicidal behaviors.

## Data Availability

All data produced in the present study are available upon reasonable request to the authors.

## Author’s contributions

Conceptualization and study design: MM and JK; first draft writing: KJ and MM; editing: all authors; recruitment of patients: JK; statistical analysis: MM and JK. All authors approved the final version of the manuscript.

## Ethics approval and consent to participate

The research project (#445/63) was approved by the Institutional Review Board of Chulalongkorn University’s institutional ethics board. All patients and controls gave written informed consent prior to participation in the study.

## Data Availability Statement

The dataset that was made and/or analysed during this study will be available from the corresponding author (MM) once it has been fully used by the authors and a reasonable request has been made.

