## Supplemental table1-2 and Figure 1-5 for "Major depression, suicidal behaviors and neuroticism are pro-atherogenic states driven by lowered reverse cholesterol transport"

### Electronic Supplementary File (ESF)

**ESF, Table 1**. Measurements of lipid profiles in subjects with metabolic syndrome (MetS) + major depression (MDD) (1.00) versus those with MetS without MDD (0.00)

**Descriptives**[a]

| | | N | Mean | Std. Deviation | Std. Error | 95% Confidence Interval for Mean Lower Bound | 95% Confidence Interval for Mean Upper Bound | Minimum | Maximum |
|---|---|---|---|---|---|---|---|---|---|
| TC | .00 | 33 | 219.091 | 50.9849 | 8.8753 | 201.012 | 237.169 | 145.0 | 322.0 |
| | 1.00 | 31 | 203.419 | 40.0930 | 7.2009 | 188.713 | 218.126 | 130.0 | 291.0 |
| | Total | 64 | 211.500 | 46.3479 | 5.7935 | 199.923 | 223.077 | 130.0 | 322.0 |
| FC | .00 | 33 | 77.9549 | 28.43466 | 4.94984 | 67.8724 | 88.0374 | 32.03 | 154.68 |
| | 1.00 | 31 | 67.2430 | 15.19735 | 2.72952 | 61.6685 | 72.8174 | 43.18 | 98.97 |
| | Total | 64 | 72.7663 | 23.44730 | 2.93091 | 66.9093 | 78.6232 | 32.03 | 154.68 |
| CE | .00 | 33 | 141.1361 | 36.12824 | 6.28912 | 128.3255 | 153.9466 | 65.42 | 206.36 |
| | 1.00 | 31 | 136.1764 | 39.77865 | 7.14446 | 121.5855 | 150.7673 | 45.91 | 202.96 |
| | Total | 64 | 138.7337 | 37.71897 | 4.71487 | 129.3118 | 148.1556 | 45.91 | 206.36 |
| CER | .00 | 33 | 64.5499 | 9.11012 | 1.58587 | 61.3196 | 67.7803 | 44.20 | 77.91 |
| | 1.00 | 31 | 65.8995 | 9.98690 | 1.79370 | 62.2363 | 69.5627 | 35.32 | 78.99 |
| | Total | 64 | 65.2036 | 9.49275 | 1.18659 | 62.8324 | 67.5749 | 35.32 | 78.99 |
| HDLc | .00 | 33 | 47.515 | 10.2839 | 1.7902 | 43.869 | 51.162 | 31.0 | 74.0 |
| | 1.00 | 31 | 48.323 | 9.1484 | 1.6431 | 44.967 | 51.678 | 31.0 | 68.0 |
| | Total | 64 | 47.906 | 9.6818 | 1.2102 | 45.488 | 50.325 | 31.0 | 74.0 |
| TG | .00 | 33 | 255.848 | 328.0081 | 57.0989 | 139.542 | 372.155 | 66.0 | 1741.0 |
| | 1.00 | 31 | 178.548 | 103.9278 | 18.6660 | 140.427 | 216.669 | 47.0 | 476.0 |
| | Total | 64 | 218.406 | 247.6044 | 30.9505 | 156.557 | 280.256 | 47.0 | 1741.0 |
| LDLc | .00 | 33 | 137.455 | 40.3013 | 7.0156 | 123.164 | 151.745 | 63.0 | 223.0 |
| | 1.00 | 31 | 136.710 | 43.7890 | 7.8647 | 120.648 | 152.772 | 44.0 | 224.0 |

| | | | | | | | | | |
|---|---|---|---|---|---|---|---|---|---|
| | Total | 64 | 137.094 | 41.6919 | 5.2115 | 126.679 | 147.508 | 44.0 | 224.0 |
| ApoA | .00 | 33 | 137.7576 | 31.01918 | 5.39975 | 126.7587 | 148.7565 | 93.00 | 224.00 |
| | 1.00 | 31 | 132.6452 | 19.07450 | 3.42588 | 125.6486 | 139.6417 | 98.00 | 172.00 |
| | Total | 64 | 135.2813 | 25.85765 | 3.23221 | 128.8222 | 141.7403 | 93.00 | 224.00 |
| ApoB | .00 | 33 | 108.6667 | 29.14690 | 5.07382 | 98.3316 | 119.0017 | 52.00 | 164.00 |
| | 1.00 | 31 | 106.3871 | 29.02605 | 5.21323 | 95.7403 | 117.0339 | 44.00 | 163.00 |
| | Total | 64 | 107.5625 | 28.87954 | 3.60994 | 100.3486 | 114.7764 | 44.00 | 164.00 |
| ApoB/ApoA | .00 | 33 | .8255 | .27050 | .04709 | .7296 | .9214 | .30 | 1.40 |
| | 1.00 | 31 | .8322 | .30204 | .05425 | .7214 | .9430 | .32 | 1.50 |
| | Total | 64 | .8287 | .28394 | .03549 | .7578 | .8997 | .30 | 1.50 |
| Castelli1 | .00 | 33 | 4.8123 | 1.55194 | .27016 | 4.2620 | 5.3626 | 2.13 | 8.70 |
| | 1.00 | 31 | 4.3879 | 1.29702 | .23295 | 3.9121 | 4.8636 | 2.04 | 6.89 |
| | Total | 64 | 4.6067 | 1.43881 | .17985 | 4.2473 | 4.9661 | 2.04 | 8.70 |
| PRO_AI | .00 | 33 | .5940052 | 1.20145105 | .20914578 | .1679892 | 1.0200212 | -1.69629 | 3.91818 |
| | 1.00 | 31 | .2564280 | .83658494 | .15025509 | -.0504338 | .5632899 | -1.63556 | 2.01097 |
| | Total | 64 | .4304912 | 1.04660670 | .13082584 | .1690567 | .6919258 | -1.69629 | 3.91818 |
| ANTI_AI or RCT | .00 | 33 | -.3911569 | .93155544 | .16216299 | -.7214721 | -.0608417 | -2.30783 | 1.71847 |
| | 1.00 | 31 | -.3840846 | .67454640 | .12115211 | -.6315102 | -.1366590 | -1.81630 | .78700 |
| | Total | 64 | -.3877312 | .81084555 | .10135569 | -.5902744 | -.1851880 | -2.30783 | 1.71847 |
| PRO/ANTI_AI | .00 | 33 | .5524374 | 1.02701711 | .17878073 | .1882730 | .9166019 | -2.00798 | 2.95574 |
| | 1.00 | 31 | .3805972 | .80692543 | .14492809 | .0846146 | .6765799 | -1.65765 | 1.73502 |
| | Total | 64 | .4692023 | .92374596 | .11546825 | .2384575 | .6999472 | -2.00798 | 2.95574 |

a. MetS_NoMetS = 1.00

**ANOVA[a]**

| | | Sum of Squares | df | Mean Square | F | Sig. |
|---|---|---|---|---|---|---|
| TC | Between Groups | 3925.724 | 1 | 3925.724 | 1.852 | .178 |
| | Within Groups | 131406.276 | 62 | 2119.456 | | |
| | Total | 135332.000 | 63 | | | |
| FC | Between Groups | 1834.124 | 1 | 1834.124 | 3.467 | .067 |
| | Within Groups | 32801.743 | 62 | 529.060 | | |
| | Total | 34635.867 | 63 | | | |
| CE | Between Groups | 393.186 | 1 | 393.186 | .273 | .603 |
| | Within Groups | 89238.221 | 62 | 1439.326 | | |
| | Total | 89631.407 | 63 | | | |
| CER | Between Groups | 29.112 | 1 | 29.112 | .320 | .574 |
| | Within Groups | 5647.967 | 62 | 91.096 | | |
| | Total | 5677.079 | 63 | | | |
| HDLc | Between Groups | 10.421 | 1 | 10.421 | .110 | .742 |
| | Within Groups | 5895.017 | 62 | 95.081 | | |
| | Total | 5905.437 | 63 | | | |
| TG | Between Groups | 95511.518 | 1 | 95511.518 | 1.572 | .215 |
| | Within Groups | 3766887.920 | 62 | 60756.257 | | |
| | Total | 3862399.437 | 63 | | | |
| LDLc | Between Groups | 8.869 | 1 | 8.869 | .005 | .944 |
| | Within Groups | 109498.569 | 62 | 1766.106 | | |
| | Total | 109507.438 | 63 | | | |
| ApoA | Between Groups | 417.780 | 1 | 417.780 | .621 | .434 |
| | Within Groups | 41705.157 | 62 | 672.664 | | |
| | Total | 42122.937 | 63 | | | |
| ApoB | Between Groups | 83.062 | 1 | 83.062 | .098 | .755 |

| | | | | | | |
|---|---|---|---|---|---|---|
| | Within Groups | 52460.688 | 62 | 846.140 | | |
| | Total | 52543.750 | 63 | | | |
| ApoB/ApoA | Between Groups | .001 | 1 | .001 | .009 | .926 |
| | Within Groups | 5.078 | 62 | .082 | | |
| | Total | 5.079 | 63 | | | |
| Castelli1 | Between Groups | 2.880 | 1 | 2.880 | 1.400 | .241 |
| | Within Groups | 127.541 | 62 | 2.057 | | |
| | Total | 130.421 | 63 | | | |
| PRO_AI | Between Groups | 1.822 | 1 | 1.822 | 1.681 | .200 |
| | Within Groups | 67.188 | 62 | 1.084 | | |
| | Total | 69.009 | 63 | | | |
| ANTI_AI or RCT | Between Groups | .001 | 1 | .001 | .001 | .973 |
| | Within Groups | 41.420 | 62 | .668 | | |
| | Total | 41.421 | 63 | | | |
| PRO/ANTI_AI | Between Groups | .472 | 1 | .472 | .549 | .461 |
| | Within Groups | 53.286 | 62 | .859 | | |
| | Total | 53.758 | 63 | | | |

a. MetS_NoMetS = 1.00

**ESF, Table 2**. Measurements of lipid profiles in subjects with metabolic syndrome (MetS) (1.00) versus healthy controls (0.00) after excluding all subjects with major depression.

**Descriptives[a]**

| | | N | Mean | Std. Deviation | Std. Error | 95% Confidence Interval for Mean | | Minimum | Maximum |
|---|---|---|---|---|---|---|---|---|---|
| | | | | | | Lower Bound | Upper Bound | | |
| TC | .00 | 34 | 189.118 | 37.9607 | 6.5102 | 175.873 | 202.363 | 107.0 | 283.0 |
| | 1.00 | 33 | 219.091 | 50.9849 | 8.8753 | 201.012 | 237.169 | 145.0 | 322.0 |
| | Total | 67 | 203.881 | 46.9979 | 5.7417 | 192.417 | 215.344 | 107.0 | 322.0 |
| FC | .00 | 34 | 52.3767 | 18.18965 | 3.11950 | 46.0300 | 58.7234 | 17.20 | 107.42 |
| | 1.00 | 33 | 77.9549 | 28.43466 | 4.94984 | 67.8724 | 88.0374 | 32.03 | 154.68 |
| | Total | 67 | 64.9749 | 26.89699 | 3.28599 | 58.4142 | 71.5356 | 17.20 | 154.68 |
| CE | .00 | 34 | 136.7410 | 33.20534 | 5.69467 | 125.1551 | 148.3268 | 83.21 | 223.74 |
| | 1.00 | 33 | 141.1361 | 36.12824 | 6.28912 | 128.3255 | 153.9466 | 65.42 | 206.36 |
| | Total | 67 | 138.9057 | 34.48256 | 4.21272 | 130.4947 | 147.3167 | 65.42 | 223.74 |
| CER | .00 | 34 | 72.3077 | 8.45500 | 1.45002 | 69.3576 | 75.2578 | 46.02 | 86.86 |
| | 1.00 | 33 | 64.5499 | 9.11012 | 1.58587 | 61.3196 | 67.7803 | 44.20 | 77.91 |
| | Total | 67 | 68.4867 | 9.55267 | 1.16704 | 66.1567 | 70.8168 | 44.20 | 86.86 |
| HDLc | .00 | 34 | 63.853 | 14.1788 | 2.4316 | 58.906 | 68.800 | 39.0 | 103.0 |
| | 1.00 | 33 | 47.515 | 10.2839 | 1.7902 | 43.869 | 51.162 | 31.0 | 74.0 |
| | Total | 67 | 55.806 | 14.8163 | 1.8101 | 52.192 | 59.420 | 31.0 | 103.0 |
| TG | .00 | 34 | 85.882 | 27.9824 | 4.7989 | 76.119 | 95.646 | 44.0 | 142.0 |
| | 1.00 | 33 | 255.848 | 328.0081 | 57.0989 | 139.542 | 372.155 | 66.0 | 1741.0 |
| | Total | 67 | 169.597 | 244.7160 | 29.8968 | 109.906 | 229.288 | 44.0 | 1741.0 |
| LDLc | .00 | 34 | 117.529 | 38.0814 | 6.5309 | 104.242 | 130.817 | 36.0 | 222.0 |
| | 1.00 | 33 | 137.455 | 40.3013 | 7.0156 | 123.164 | 151.745 | 63.0 | 223.0 |
| | Total | 67 | 127.343 | 40.1662 | 4.9071 | 117.546 | 137.141 | 36.0 | 223.0 |

| | | | | | | | | | |
|---|---|---|---|---|---|---|---|---|---|
| ApoA | .00 | 34 | 150.0588 | 30.29946 | 5.19631 | 139.4868 | 160.6308 | 96.00 | 231.00 |
| | 1.00 | 33 | 137.7576 | 31.01918 | 5.39975 | 126.7587 | 148.7565 | 93.00 | 224.00 |
| | Total | 67 | 144.0000 | 31.04737 | 3.79304 | 136.4270 | 151.5730 | 93.00 | 231.00 |
| ApoB | .00 | 34 | 80.6176 | 21.00507 | 3.60234 | 73.2886 | 87.9467 | 28.00 | 131.00 |
| | 1.00 | 33 | 108.6667 | 29.14690 | 5.07382 | 98.3316 | 119.0017 | 52.00 | 164.00 |
| | Total | 67 | 94.4328 | 28.84663 | 3.52418 | 87.3966 | 101.4691 | 28.00 | 164.00 |
| ApoB/ApoA | .00 | 34 | .5548 | .17510 | .03003 | .4937 | .6159 | .22 | .99 |
| | 1.00 | 33 | .8255 | .27050 | .04709 | .7296 | .9214 | .30 | 1.40 |
| | Total | 67 | .6881 | .26345 | .03218 | .6239 | .7524 | .22 | 1.40 |
| Castelli1 | .00 | 34 | 3.0542 | .73925 | .12678 | 2.7962 | 3.3121 | 1.98 | 4.88 |
| | 1.00 | 33 | 4.8123 | 1.55194 | .27016 | 4.2620 | 5.3626 | 2.13 | 8.70 |
| | Total | 67 | 3.9201 | 1.49175 | .18225 | 3.5563 | 4.2840 | 1.98 | 8.70 |
| PRO_AI | .00 | 34 | -.6281470 | .79524557 | .13638349 | -.9056213 | -.3506727 | -2.49869 | 1.09066 |
| | 1.00 | 33 | .5940052 | 1.20145105 | .20914578 | .1679892 | 1.0200212 | -1.69629 | 3.91818 |
| | Total | 67 | -.0261914 | 1.18112930 | .14429791 | -.3142915 | .2619086 | -2.49869 | 3.91818 |
| ANTI_AI or RCT | .00 | 34 | .6481589 | 1.00271406 | .17196404 | .2982954 | .9980223 | -1.50503 | 2.83939 |
| | 1.00 | 33 | -.3911569 | .93155544 | .16216299 | -.7214721 | -.0608417 | -2.30783 | 1.71847 |
| | Total | 67 | .1362571 | 1.09432257 | .13369278 | -.1306692 | .4031833 | -2.30783 | 2.83939 |
| PRO/ANTI_AI | .00 | 34 | -.7644494 | .85176689 | .14607682 | -1.0616449 | -.4672539 | -2.63196 | .79805 |
| | 1.00 | 33 | .5524374 | 1.02701711 | .17878073 | .1882730 | .9166019 | -2.00798 | 2.95574 |
| | Total | 67 | -.1158335 | 1.14637377 | .14005185 | -.3954560 | .1637890 | -2.63196 | 2.95574 |

a. MDD_NoMDD = .00

**ANOVA[a]**

| | | Sum of Squares | df | Mean Square | F | Sig. |
|---|---|---|---|---|---|---|

| | | | | | | |
|---|---|---|---|---|---|---|
| TC | Between Groups | 15044.788 | 1 | 15044.788 | 7.480 | .008 |
| | Within Groups | 130736.257 | 65 | 2011.327 | | |
| | Total | 145781.045 | 66 | | | |
| FC | Between Groups | 10956.121 | 1 | 10956.121 | 19.356 | <.001 |
| | Within Groups | 36791.458 | 65 | 566.022 | | |
| | Total | 47747.580 | 66 | | | |
| CE | Between Groups | 323.486 | 1 | 323.486 | .269 | .606 |
| | Within Groups | 78153.611 | 65 | 1202.363 | | |
| | Total | 78477.096 | 66 | | | |
| CER | Between Groups | 1007.845 | 1 | 1007.845 | 13.063 | <.001 |
| | Within Groups | 5014.893 | 65 | 77.152 | | |
| | Total | 6022.737 | 66 | | | |
| HDLc | Between Groups | 4469.970 | 1 | 4469.970 | 29.001 | <.001 |
| | Within Groups | 10018.507 | 65 | 154.131 | | |
| | Total | 14488.478 | 66 | | | |
| TG | Between Groups | 483774.348 | 1 | 483774.348 | 9.065 | .004 |
| | Within Groups | 3468697.772 | 65 | 53364.581 | | |
| | Total | 3952472.119 | 66 | | | |
| LDLc | Between Groups | 6648.452 | 1 | 6648.452 | 4.329 | .041 |
| | Within Groups | 99830.652 | 65 | 1535.856 | | |
| | Total | 106479.104 | 66 | | | |
| ApoA | Between Groups | 2534.057 | 1 | 2534.057 | 2.696 | .105 |
| | Within Groups | 61085.943 | 65 | 939.784 | | |
| | Total | 63620.000 | 66 | | | |
| ApoB | Between Groups | 13175.085 | 1 | 13175.085 | 20.514 | <.001 |
| | Within Groups | 41745.363 | 65 | 642.236 | | |

| | | | | | | |
|---|---|---|---|---|---|---|
| | Total | 54920.448 | 66 | | | |
| ApoB/ApoA | Between Groups | 1.227 | 1 | 1.227 | 23.793 | <.001 |
| | Within Groups | 3.353 | 65 | .052 | | |
| | Total | 4.581 | 66 | | | |
| Castelli1 | Between Groups | 51.764 | 1 | 51.764 | 35.378 | <.001 |
| | Within Groups | 95.107 | 65 | 1.463 | | |
| | Total | 146.871 | 66 | | | |
| PRO_AI | Between Groups | 25.013 | 1 | 25.013 | 24.244 | <.001 |
| | Within Groups | 67.061 | 65 | 1.032 | | |
| | Total | 92.074 | 66 | | | |
| ANTI_AI or RCT | Between Groups | 18.089 | 1 | 18.089 | 19.291 | <.001 |
| | Within Groups | 60.949 | 65 | .938 | | |
| | Total | 79.038 | 66 | | | |
| PRO/ANTI_AI | Between Groups | 29.041 | 1 | 29.041 | 32.719 | <.001 |
| | Within Groups | 57.694 | 65 | .888 | | |
| | Total | 86.735 | 66 | | | |

a. MDD_NoMDD = .00

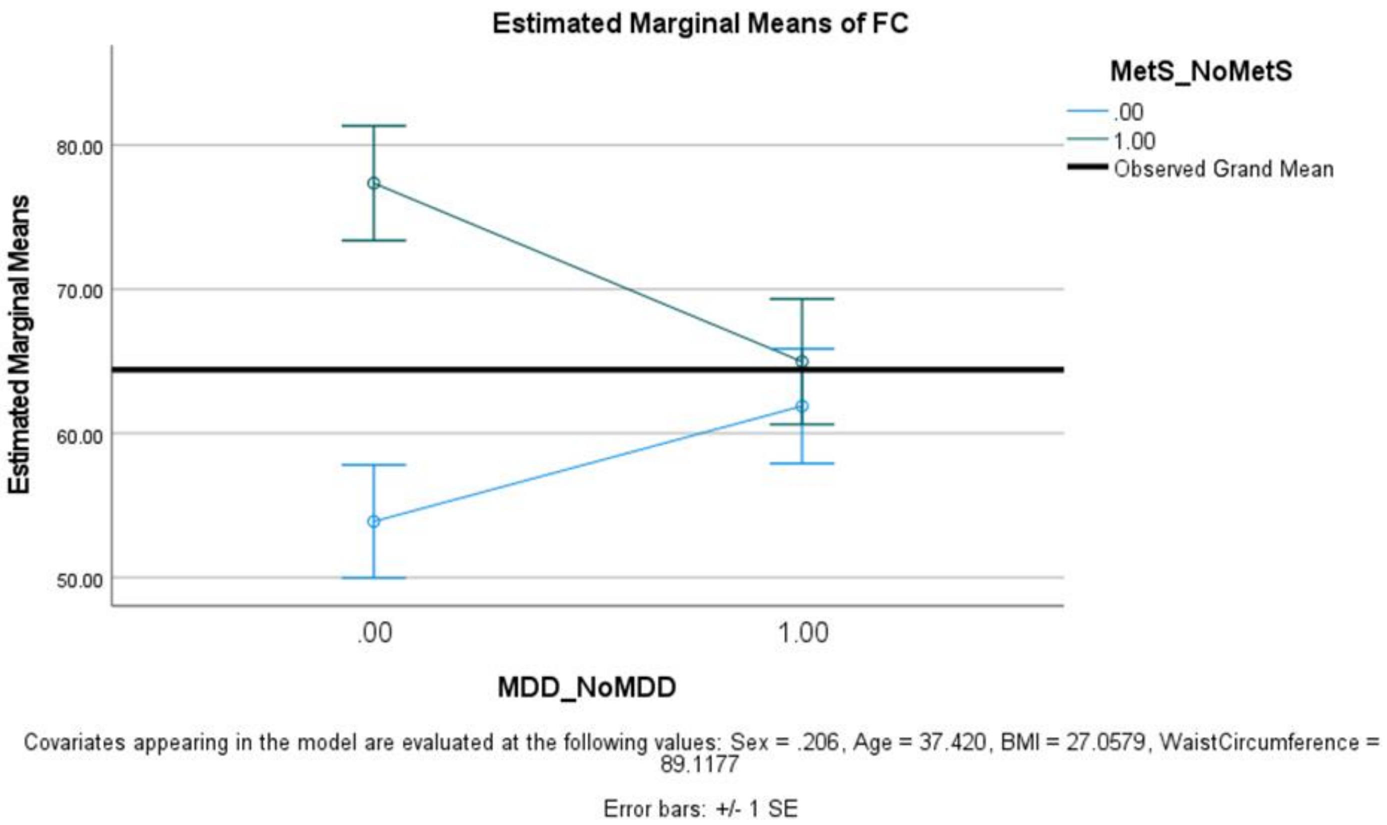


**ESF, Figure 1.** Interaction pattern between major depression (MDD versus healthy controls, HC) and metabolic syndrome (MetS). Shown are the estimated marginal mean values of free cholesterol (FC) after covarying for age, sex, body mass index and waist circumference (0.00=controls; 1.00=MDD).

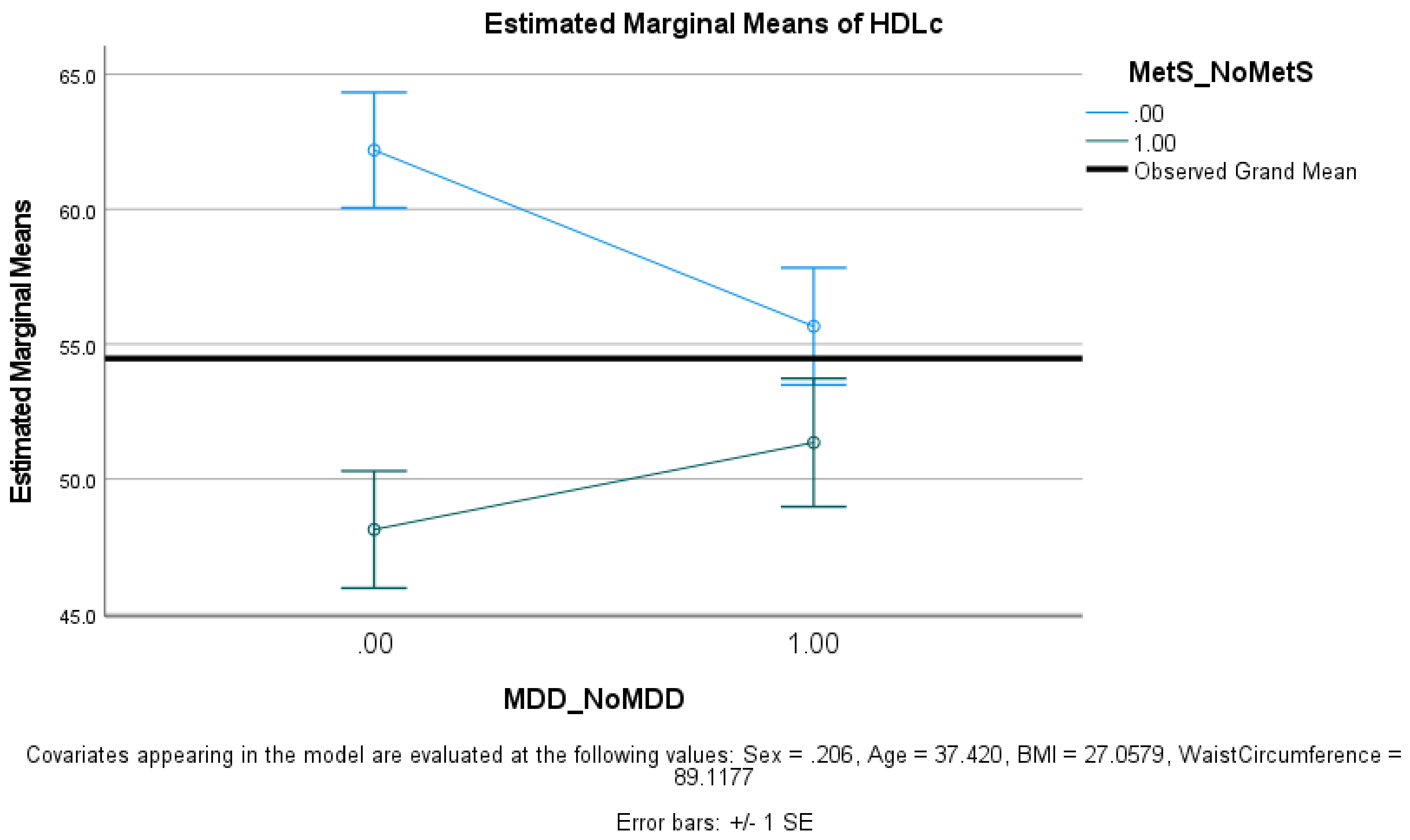


**ESF, Figure 2.** Interaction pattern between major depression (MDD versus healthy controls, HC) and metabolic syndrome (MetS). Shown are the estimated marginal mean values of high-density lipoprotein cholesterol (HDLc) after covarying for age, sex, body mass index and waist circumference (0.00=controls; 1.00=MDD).

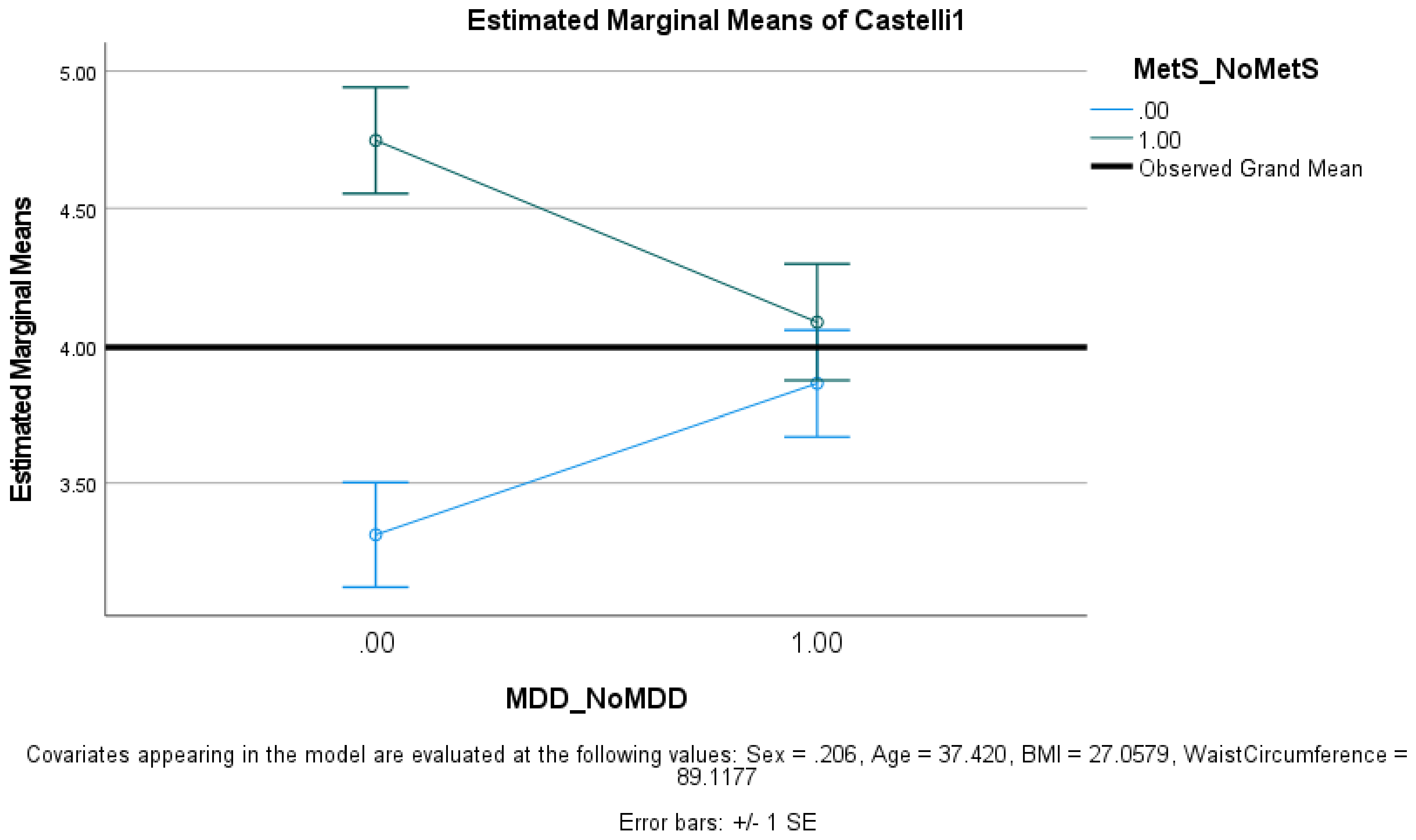


**ESF, Figure 3.** Interaction pattern between major depression (MDD versus healthy controls, HC) and metabolic syndrome (MetS). Shown are the estimated marginal mean values of the Castelli risk index 1 (0.00=controls; 1.00=MDD).

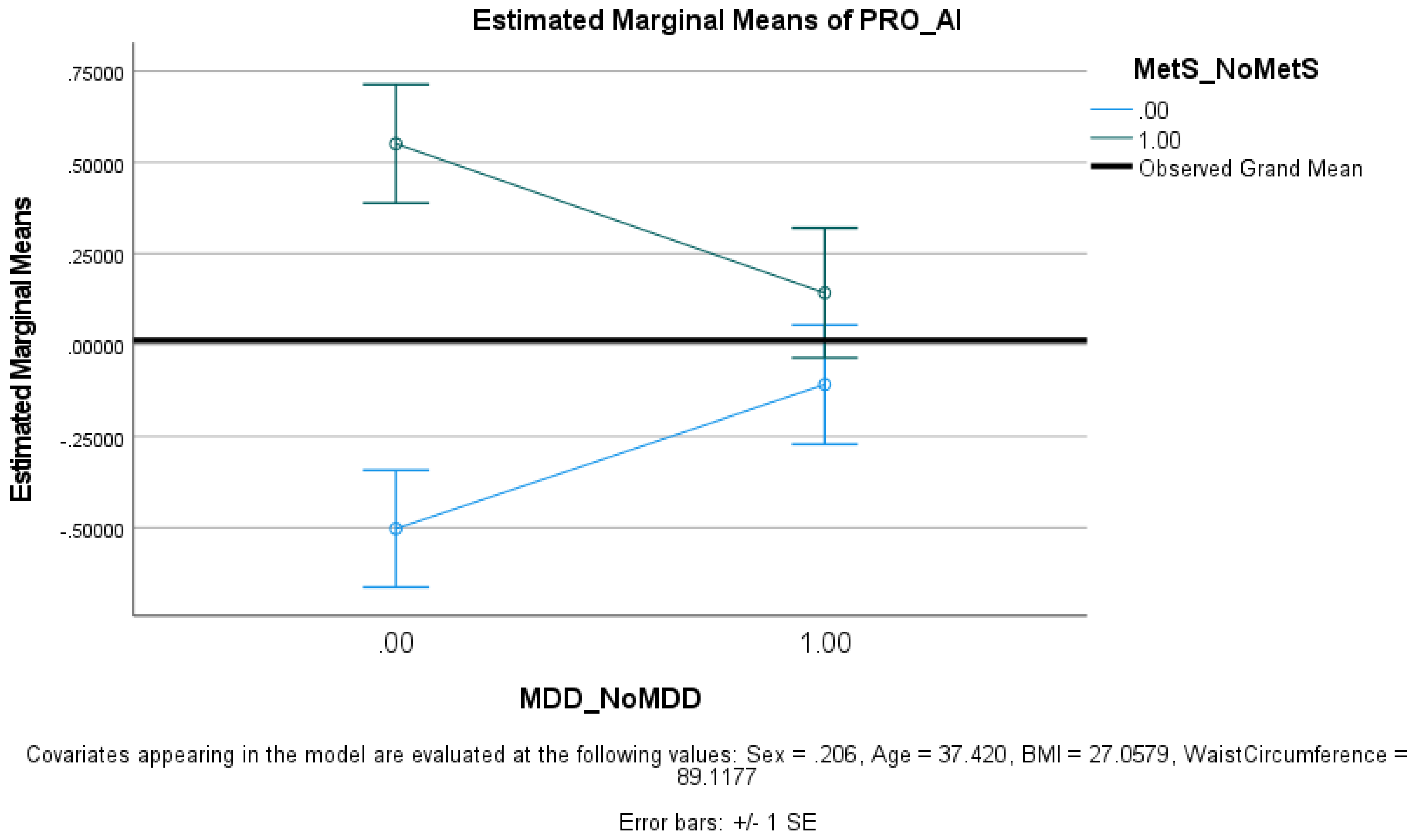


**ESF, Figure 4.** Interaction pattern between major depression (MDD versus healthy controls, HC) and metabolic syndrome (MetS). Shown are the estimated marginal mean values of the pro-atherogenic index (PRO_AI) (0.00=controls; 1.00=MDD).

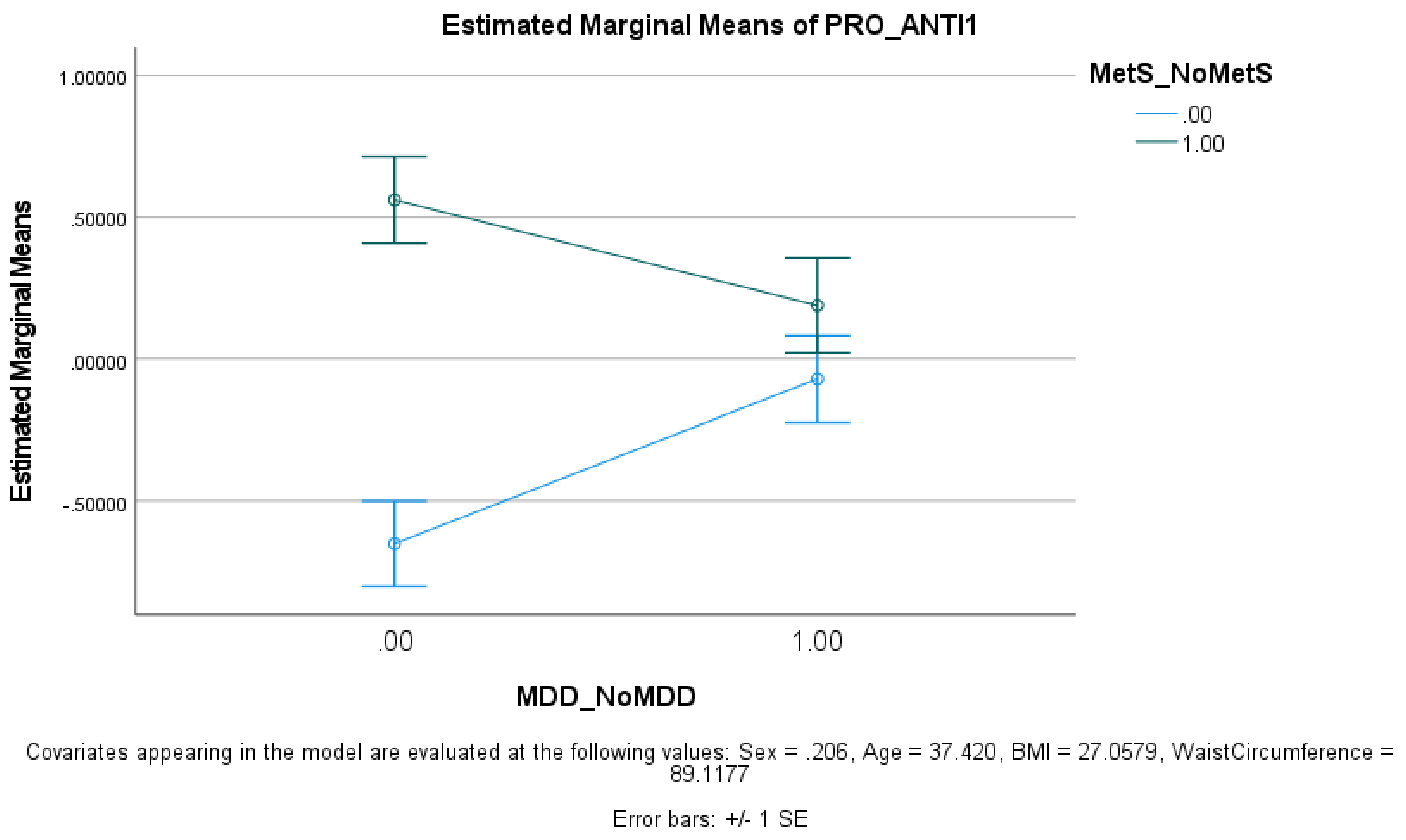


**ESF, Figure 5.** Interaction pattern between major depression (MDD versus healthy controls, HC) and metabolic syndrome (MetS). Shown are the estimated marginal mean values of the pro- versus anti-atherogenic index (PRO/ANTI_AI) (0.00=controls; 1.00=MDD).
